# Mapping abstraction and metacognition onto distinct transdiagnostic symptom profiles

**DOI:** 10.64898/2026.06.17.26354096

**Authors:** Taiki Oka, Yoshihiko Kunisato, Kotaro Koizumi, Misa Murakami, Hugo Six, Jessica E Taylor, Aurelio Cortese

## Abstract

Transdiagnostic psychiatric research on reward-guided learning has largely focused on simple associative processes, leaving it unclear whether or how higher-level processes are disrupted. Here, we studied how abstraction, the ability to extract relevant features from complex information, and metacognition, the ability to monitor and evaluate one’s own mental processes, map onto specific transdiagnostic dimensions. Using an online sample (N = 249), we examined associations between these processes and three cross-culturally robust transdiagnostic dimensions derived from a large existing dataset (N = 19,505): Compulsive hypersensitivity, Social withdrawal, and Addictive behaviours. Computational modelling of an abstract representation learning task with confidence judgments revealed that Compulsive hypersensitivity was negatively associated with both abstraction ability (*p_boot_* = 0.003) and metacognitive sensitivity (*p_boot_* = 0.005), while Social withdrawal was positively associated with metacognitive sensitivity alone (*p_boot_* = 0.002). Moreover, transdiagnostic dimensions revealed more coherent associations with higher-order cognition than symptom-level analyses, highlighting the added value of examining psychopathology at the factor rather than the symptom level. These findings portray a hierarchical view of cognitive dysfunctions in psychopathology and point to representational and metacognitive processes as potential targets for transdiagnostic intervention.

## Introduction

Impairments in reward-guided learning are widely observed across psychiatric disorders and are considered a hallmark of psychopathology (Maia and Frank, 2011). While a growing number of studies have adopted transdiagnostic approaches to investigate shared mechanisms across disorders (Fried, 2022; Kist et al., 2023), this body of work has largely focused on relatively simple learning processes (Lee et al., 2025; Wise et al., 2023). Whether these findings extend to the broader cognitive architecture that supports learning in more complex scenarios remains an open question.

Here, we investigate two such candidate mechanisms — abstraction and metacognition — and examine how each is associated with transdiagnostic symptom dimensions. Abstraction is defined as the process of mentally extracting and organising, from complex information, a small set of features relevant to the task or context at hand (De Martino and Cortese, 2022; Ho et al., 2019), and is critical for generalising behaviour across diverse scenarios. Metacognition, in turn, monitors and evaluates these abstract representations (De Martino and Cortese, 2022; Lee and Hare, 2023). Both processes have been independently implicated in psychopathology. Impairments in metacognition have been reported across multiple transdiagnostic studies (Katyal et al., 2025; Seow et al., 2025a; T. Seow et al., 2021), while abstraction deficits have been documented in specific psychiatric conditions (Gjelsvik et al., 2018; Kobayashi et al., 2014). However, no previous work has examined them concomitantly, nor has it examined how they might be associated with transdiagnostic dimensions. This gap exists partly because abstraction is an inherently internal and latent process that previous studies have assessed using self-reported or behavioural measures alone, which cannot directly capture latent computational mechanisms (Eling et al., 2008). Additionally, although metacognition has been studied from a transdiagnostic perspective (Hoven et al., 2023), abstraction and metacognition have, to the best of our knowledge, not been measured and characterised within the same transdiagnostic framework and the same experiment.

Addressing this question requires both a principled computational approach to quantifying abstract processes and a transdiagnostic framework (Gillan and Seow, 2020; Wise et al., 2023). In this study, we defined abstraction as an algorithmic strategy in an associative learning task in which participants learned to identify which subset of stimulus features predicted reward. The task effectively required participants to extract a low-dimensional task-relevant structure from higher-dimensional information. We then used a computational model to derive a continuous abstraction metric at the individual level, alongside measures of metacognition based on subjective confidence ratings. These were examined across participants in relation to transdiagnostic psychiatric symptom dimension scores obtained by decomposing co-occurring psychiatric symptoms across a large online sample into a small set of latent factors, each capturing shared variance across diagnostic boundaries (Wise et al., 2023). These dimensions were derived by applying factor weights computed from an existing large-scale study (N = 19,505) to psychiatric questionnaire scores of our sample (N = 249). This computational factor modelling approach allowed us to ask how transdiagnostic symptom dimensions relate to abstraction and metacognition (Figure 1).

**Figure 1:**
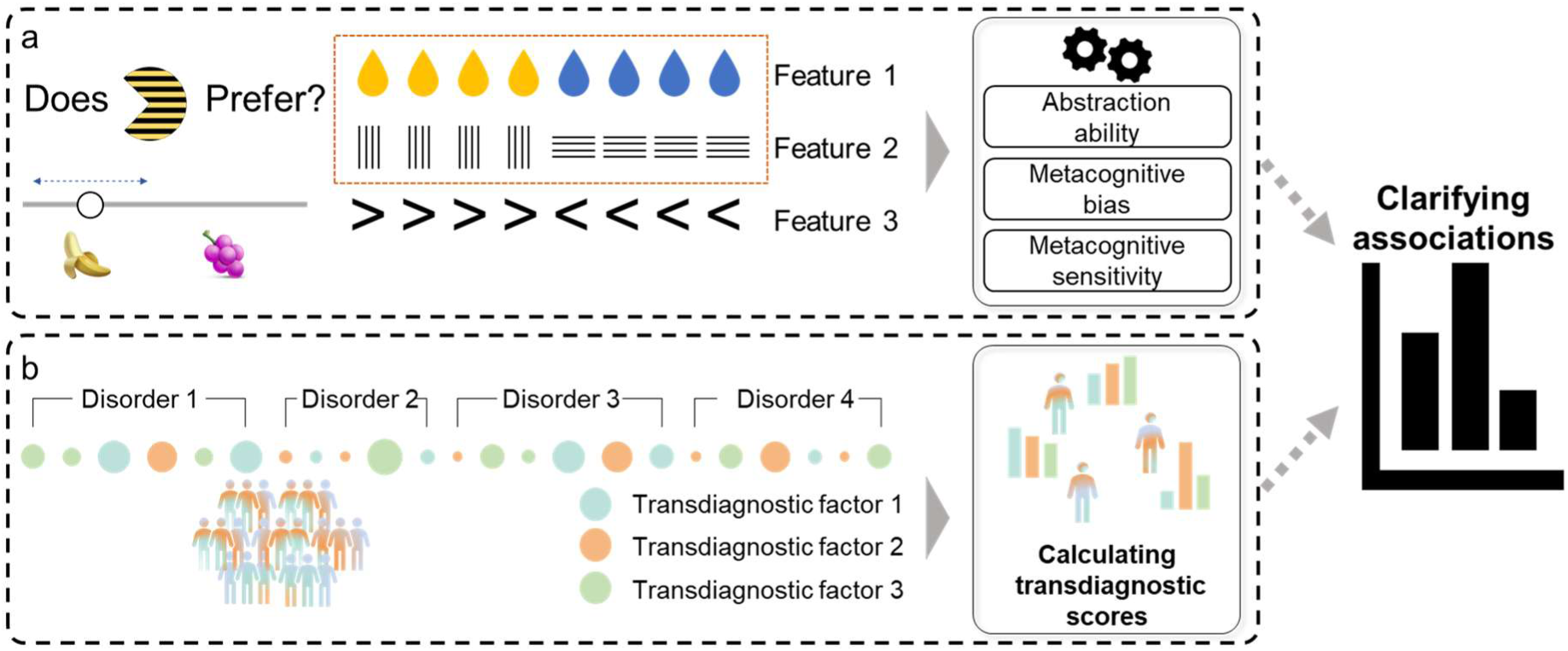
Computational factor modelling framework and behavioural task. Computational factor modelling approach: **a.** Abstract representation learning task: Participants learned the fruit preferences of Pac-Man-like characters, with the features that predict preference changing block-by-block. Three types of feature combinations could predict preference: colour–stripe orientation, colour–mouth direction, and stripe orientation–mouth direction (see the example in the top-right box). The cursor on the slider allows participants to report both their choice and their confidence simultaneously. Latent behavioural scores are estimated through computational modelling. **b.** Transdiagnostic psychiatric dimension scores are extracted across diagnostic boundaries constructed by factor analysis. Then, extracted latent behavioural variables can be linked to each transdiagnostic psychiatric dimension. This data-driven approach enables an integrated understanding of how computational mechanisms underlying key behaviour attributes map onto transdiagnostic symptom dimensions.

## Results

### Behavioural validation of an abstract representation learning task

We recruited participants from the general population worldwide (N = 512) via an online experimental platform (Prolific, https://www.prolific.co/) and analysed 249 participants after applying pre-registered exclusion criteria (e.g., failing attention checks, performing the task at chance level; see Methods for further details, and Supplementary Methods for demographics and discussion on sample characteristics). Participants completed self-report questionnaires (consisting of multiple symptom rating scales--8 questionnaires, 176 items), including those related to several types of addictions and to internalising disorders such as social anxiety, attention deficit hyperactivity disorder, etc., see Methods for further details) and a behavioural task. In the behavioural experiment, participants had to learn the preferred fruit of a Pac-Man-like character (Figure 1a). These characters comprised three features: colour (blue/yellow), stripe orientation (vertical/horizontal), and mouth direction (right/left). Fruit preference was determined by only two of these features at a time, with preferences randomly changing in each new block (see Supplementary Figure 2 for the details). Participants indicated their choice (selecting the character’s fruit preference) as well as their confidence (e.g., more lateral position = higher confidence) using a cursor (see Figure 1a; reaction time and confidence distribution are shown in Supplementary Figures 2 and 3). The cursor’s randomised starting position only marginally affected the final response (see Supplementary Figure 4).

We confirmed learning curves within blocks (Figure 2a) and across blocks (Figure 2b), observing a positive relationship between learning speed and time, although the latter effect did not reach statistical significance (*β =* 0.0058*, p* = 0.13; see Supplementary materials for model details). Learning speed, however, correlated with overall confidence (*β =* 0.026, *p* = 0.008; see Figure 2c). These results are consistent with our prior findings (Cortese et al., 2021), indicating that this online experiment is valid relative to our original lab-based experiment.

**Figure 2:**
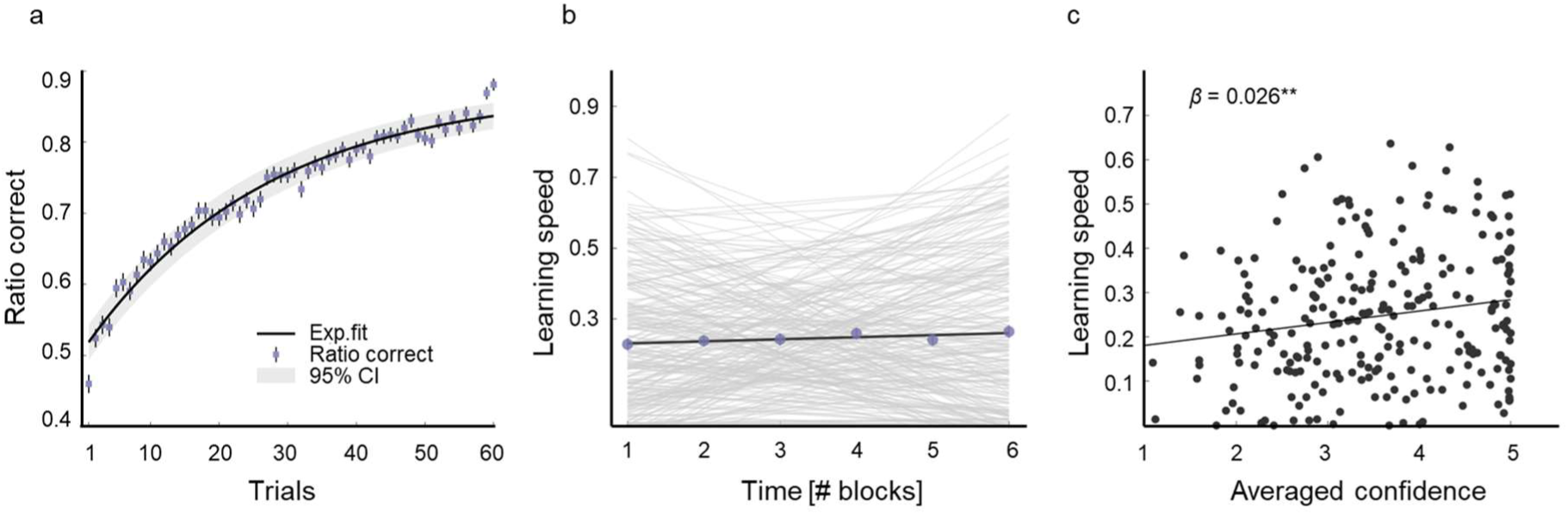
Behaviour validation of the abstract representation learning task. **a.** Trial-by-trial ratio of correct responses served as an index of within-block learning. Each dot represents the mean across participants, error bars denote the standard error of the mean (SEM), and the shaded area indicates the 95% confidence interval. Ratio-correct values were computed for each trial between completed blocks and then averaged across participants. **b.** Relationship between learning speed and time across participants. Learning speed was defined as the inverse of the maximum-normalised number of trials required to complete a block. Thin grey lines show individual participants’ data, and the bold black line represents the group-average fit. Circles represent population-level mean values, and error bars represent the SEM. **c.** Averaged confidence ratings were positively associated with learning speed across participants. Each point corresponds to a single participant, and the solid line represents the regression fit. ** p < 0.01.

### Extraction and validation of transdiagnostic psychiatric dimensions

To extract transdiagnostic dimensional scores, we transformed initial scores from a battery of psychiatric questionnaires (see Methods) using weights derived from a previous large-scale study (from the Japanese population, N = 19,505) (Oka et al., 2025), in line with previous methods (Seow et al., 2025a; Tricia Seow et al., 2021). We favoured this approach because our sample size (249) was limited relative to the total number of questionnaire items (176), resulting in an insufficient sample-to-item ratio for reliable de novo factor analysis (Osborne and Costello, 2004). The correlations between the questionnaires are shown in Supplementary Figure 5.

Nevertheless, we still sought to assess the validity of using the large-sample factor weights. To this end, we conducted a de novo exploratory factor analysis on the newly collected sample, identified three transdiagnostic factors, and compared the results with the original weights from the previous study. All factors showed significant correlations with moderate effect sizes (*r* > 0.4), indicating replication of the same factor structure and its cross-cultural generalisation (Factor 1: Spearman’s *r* = 0.63; Factor 2: Spearman’s *r* = 0.44; Factor 3: Spearman’s *r* = 0.46, see Figure 3). To maintain consistency with previous work, we refer to these dimensions as follows (Oka et al., 2025). *Compulsive hypersensitivity* is defined primarily by obsessive-compulsive disorder and attention-deficit/hyperactivity disorder symptomatology, reflecting a transdiagnostic tendency toward compulsive and impulsive dyscontrol. *Social withdrawal* is characterised predominantly by social anxiety and psychological distress, capturing a disposition toward avoidance of social interaction. *Addictive behaviours* is defined largely by gaming disorder, alcohol-related problems, and nicotine dependence, reflecting a shared propensity toward behavioural and substance-related addiction.

**Figure 3.**
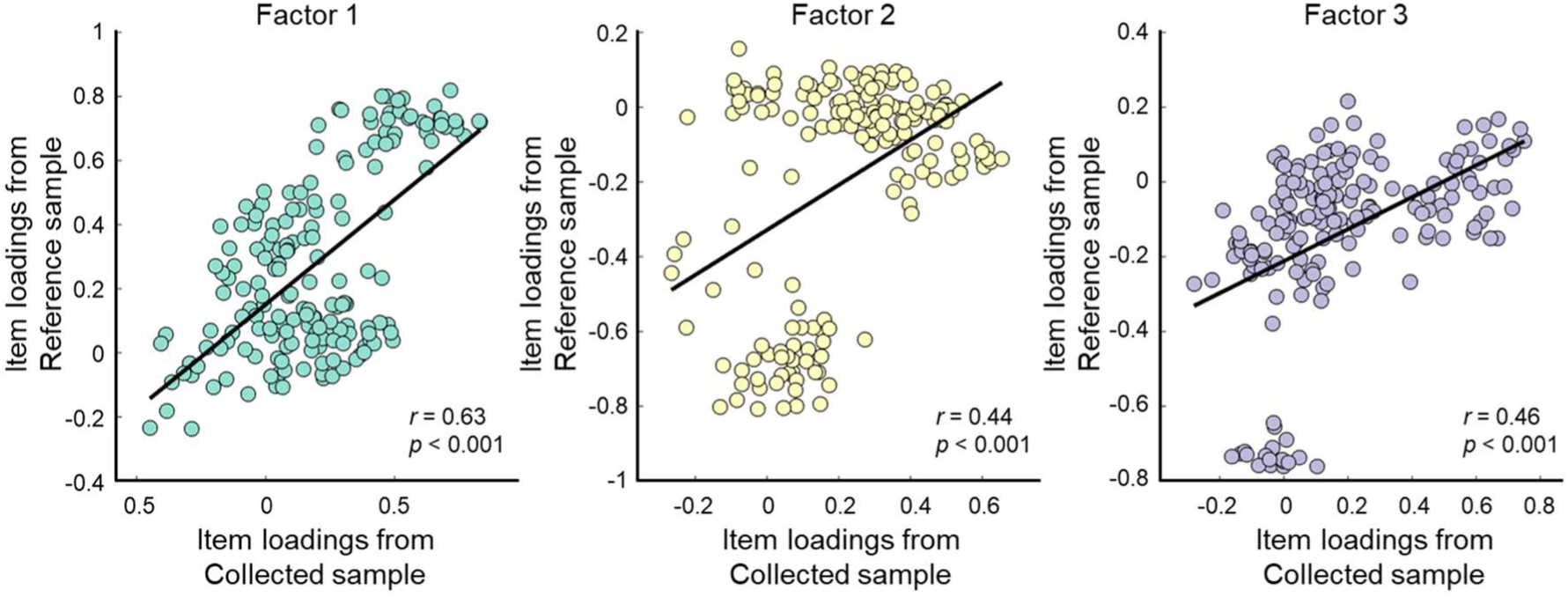
Correlations between item loadings for the three-factor structure from de-novo factor analysis between a collected sample in this study (N = 249) and a larger reference dataset (N = 19,505). Scatter plot of weights of each transdiagnostic factor between the newly collected global sample (from this study, x-axis) and the reference sample (from a large online survey in Japan(Oka et al., 2025), y-axis). Each circle indicates a single questionnaire item. Correlation values are as follows: FA1 r = 0.63, 95% CI [0.53, 0.71], p < 0.001. FA2 r = 0.44, 95% CI [0.31, 0.55], p < 0.001, FA3: r = 0.46, 95% CI [0.34, 0.57], p < 0.001.

### Abstraction is associated with the Compulsive hypersensitivity dimension, extending beyond findings for individual symptoms

We first quantified individual differences in abstraction by comparing two reinforcement learning models that differed in their feature-level representations (see Methods and the original paper (Cortese et al., 2021)). In short, the “Abstract RL” model assumes that participants rely on a reduced, lower-dimensional representation of task features, whereas the “Feature RL” model assumes that participants learn the full feature combination (Figure 4a, b). We used Hierarchical Bayesian estimation (Piray et al., 2019) to perform concurrent parameter fitting (learning rate *α* and inverse temperature *β*) and model comparison across the two RL strategies in each block. The participant count for the best-fitting model in each block is shown in Supplementary Figure 6. Both parameters showed adequate recovery (*α*: Spearman’s *r* = 0.40, *β*: Spearman’s *r* = 0.72, both *p* < .001; Supplementary Figure 7, see also Methods for details on parameter fitting and recovery procedures). We quantified our abstraction ability metric as the mean posterior responsibility of the Abstract RL model across blocks within each participant, as computed through HBI. This continuous metric reflects the degree to which each participant’s behaviour was better explained by abstract rather than feature-level representations across the task. Although our preregistration specified the proportion of blocks in which the Abstract RL model provided a better fit as the abstraction metric, we deviated from this approach because the proportion measure takes only seven discrete values (6 blocks per participant), resulting in limited variance and reduced sensitivity to individual differences. The continuous responsibility metric retains the same theoretical interpretation while providing greater resolution (see the histogram of abstract RL use probability in Figure 4c).

**Figure 4.**
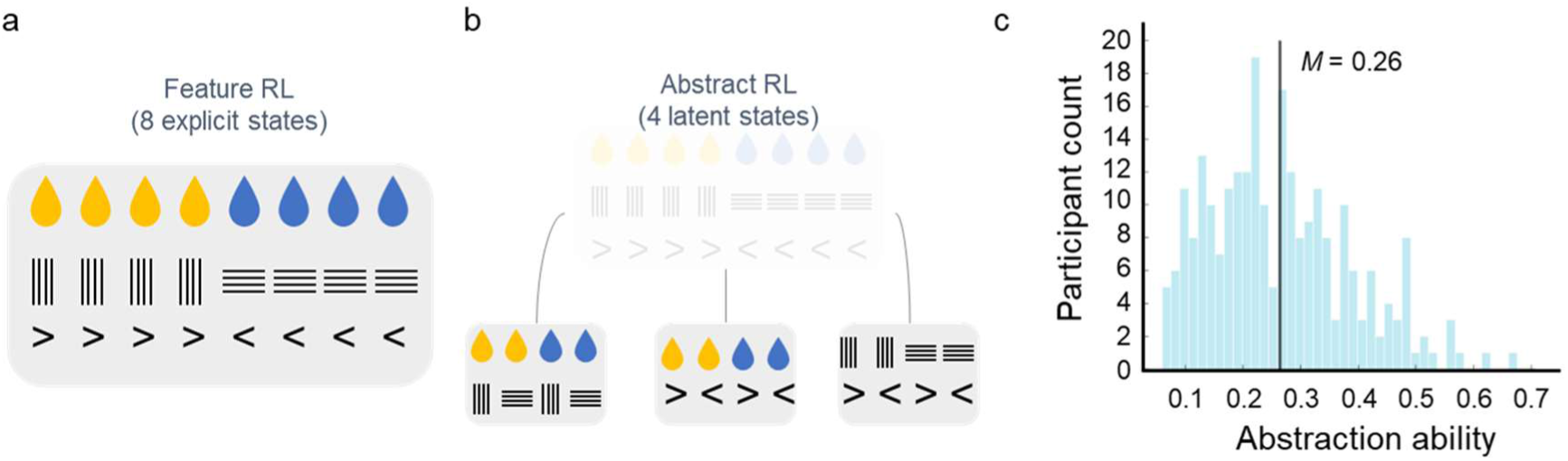
Behaviour modelling framework. **a.** Feature RL model. The model has 2^3^ = 8 states, each corresponding to a combination of all three features. **b.** Abstract RL model. The model is designed with 2^2^ = 4 states, where each state is defined by the combination of two features, with three possible ways to achieve this 4-state configuration. **c.** Histogram of probability of using the abstract strategy across participants. The vertical line shows the population mean (M = 0.26).

We then examined the association between the abstraction metric estimated from computational modelling and psychopathological symptoms, controlling for sex, age, educational level, and processing speed [see Methods for details]. Symptom-level and transdiagnostic dimension-level models were tested independently, with the expectation that transdiagnostic dimensions would reveal more specific associations than individual symptom scores, reflecting the added value of a transdiagnostic approach over symptom-by-symptom analysis. This two-level approach was applied consistently across all subsequent regression analyses. To test these relationships, we employed bootstrap multiple linear regression (5,000 iterations with replacement), reporting the mean beta estimate, 95% confidence interval (2.5th and 97.5th percentiles of the bootstrap distribution), and a two-tailed bootstrap p-value. Although our pre-registered analysis plan specified standard multiple linear regression, we deviated from this protocol in favour of bootstrap regression to provide more robust inference without distributional assumptions (pre-registered regression analyses are reported in Supplementary Figures 8-11 and Supplementary Tables 1-4, which were almost consistent with the bootstrap results below. In particular, results using the preregistered abstraction metric based on proportion of best-fit models across blocks are reported in Supplementary Figure 9 and Supplementary Table 2 for transparency.) All statistical values for all analyses are shown in Supplementary Tables 5-7. Bootstrapping results showed negative relationships between abstraction ability and ADHD, psychological distress, and obsessive-compulsive symptoms (*β* = −0.017 [−0.030 −0.004], *p_boot_* = 0.032; *β* = −0.020 [−0.034 −0.006], *p_boot_* = 0.024; *β* = −0.026 [−0.040 −0.012], *p_boot_* = 0.003, respectively; see Supplementary Figure 12c). Note that these *ps_boot_* are adjusted for multiple comparisons (corrected for false discovery rate (FDR)). Importantly, we found a significantly negative relationship with Compulsive hypersensitivity within the transdiagnostic dimensions (*β_boot_* = −0.037 [−0.061 −0.012], *p_boot_* = 0.003, see Figure 5a). Notably, not all symptoms with high loadings on the Compulsive hypersensitivity factor showed significant associations at the symptom level, suggesting that the transdiagnostic dimension may capture shared variance extending beyond individual symptom scores.

**Figure 5.**
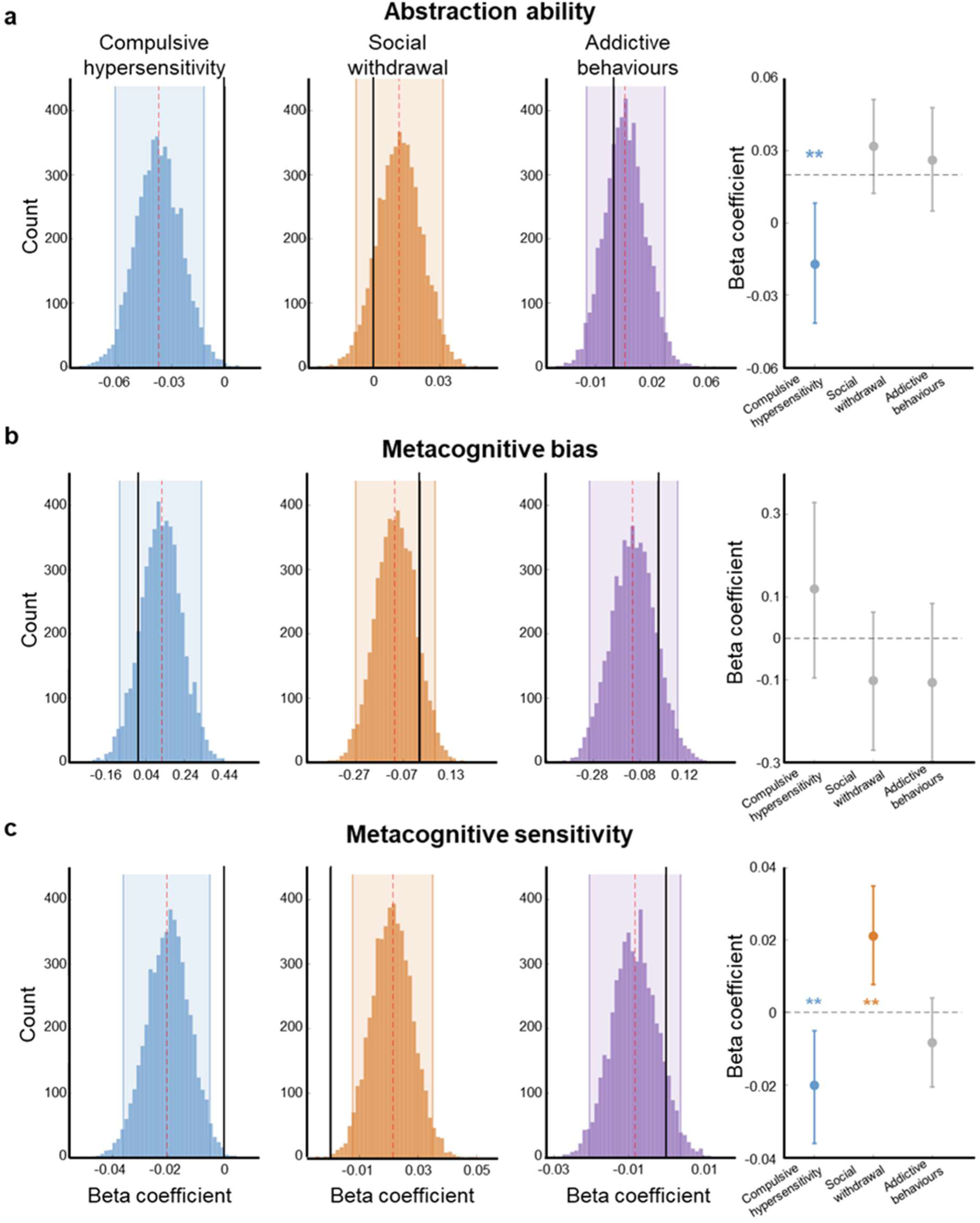
Associations between task-related behaviour measures and transdiagnostic symptom dimensions. Regression coefficients between each metric and dimension scores based on bootstrap multiple linear regression estimation with 5000 iterations. **a.** Associations between the abstraction parameter and each dimension score. **b.** Associations between the averaged confidence and each dimension score**. c.** Associations between the metacognitive sensitivity and each dimension score. The black vertical line in each histogram represents the hypothetical null effect (coefficient = 0). The red vertical dashed line in each histogram represents the average beta coefficient. ** p_boot_ < 0.01

### Transdiagnostic dimensions are not associated with metacognitive bias

We next conducted a pre-registered analysis using the same method, except that we included participants’ response bias as a covariate (Sarna et al., 2026). We first tested the association between the averaged confidence and psychopathology. If confidence was biased towards high (or low) ratings, it should reflect a metacognitive bias, defined as the degree to which the self-rating diverges directionally from reality, resulting in under- or over-confidence (Seow et al., 2025a). Our results indicated no statistically significant relationships between averaged confidence (metacognitive bias) and psychopathology after correcting for multiple comparisons (note that several associations between metacognitive bias and symptom-level scores were significant before correction, Figure 5b, Supplementary Figure 12b).

### Transdiagnostic dimensions are associated with metacognitive sensitivity

Finally, we tested the relationship between psychopathology and metacognitive sensitivity. Simply speaking, this measure captures how well confidence ratings predict choice accuracy, and we measured it as the area under the type 2 ROC curve (AUROC2) (Embon et al., 2023; Jia et al., 2020; Pinkham et al., 2018). Results showed negative relationships between metacognitive sensitivity and obsessive-compulsive (*β* = −0.017 [−0.025 −0.008], *p_boot_* = 0.004) and gaming disorder symptoms (*β* = −0.009 [−0.020 −0.003], *p_boot_* = 0.032). Note that these *p_boot_* are adjusted for multiple comparisons (See Supplementary Figure 12c). Transdiagnostically, we found a significantly negative relationship with Compulsive hypersensitivity (*β_boot_* = −0.020 [−0.036 −0.005], *p_boot_* = 0.005) and a positive relationship with Social withdrawal dimensions (*β_boot_* = 0.021 [0.008 0.035], *p_boot_* = 0.002, see Figure 5c). Models based on single multiple linear regression that included the interaction between psychopathology and metacognitive sensitivity did not provide a better fit to the data, as indicated by a likelihood-ratio test (all *ps* > 0.05, Supplementary Table 8).

## Discussion

In this preregistered study, we examined the relationship between abstraction, metacognition, and transdiagnostic psychopathology in an online sample. First, we found that transdiagnostic symptom dimensions appear broadly consistent across samples (even across countries). Our computationally derived metric of abstraction ability was associated with Compulsive hypersensitivity, beyond what was observed across symptoms that collectively define this dimension. Additionally, task-related metacognitive sensitivity, extracted from confidence judgments and choice data, was distinctly associated with transdiagnostic dimensions. The Social withdrawal symptom dimension was associated with heightened metacognitive sensitivity, while the Compulsive hypersensitivity symptom dimension was associated with reduced metacognitive sensitivity. Taken together, our results indicate multilayered effects of psychopathology across both abstract task representation and metacognitive evaluation.

The selective association between reduced abstraction and the Compulsive hypersensitivity dimension provides important mechanistic insight at the transdiagnostic level. Difficulty compressing task-relevant information into lower-dimensional representations may reflect an over-reliance on detailed, feature-level encoding, consistent with the inflexible, stimulus-bound behavioural control characteristic of compulsive psychopathology (Robbins et al., 2024). Our findings are also consistent with previous converging evidence linking compulsive tendencies to less efficient computation and a preference for familiar over goal-directed action (Banca et al., 2024; Doyle et al., 2026). Alternatively, this need not necessarily imply an impaired capacity to form abstractions *per se*, but may instead reflect a tendency to perseverate within existing representational structures even when more flexible updating would be adaptive (Tricia Seow et al., 2021). However, existing evidence in compulsive psychopathology is inconsistent (Benzina et al., 2021), making it difficult to draw firm conclusions in favour of a perseveration account. Future work directly contrasting the formation of new abstractions with the flexible updating of existing ones will be needed to arbitrate between these accounts. That these associations were not observed for Social withdrawal or Addictive behaviours suggests that impaired abstraction is a dimension-specific marker of the compulsive spectrum rather than a transdiagnostic feature of psychopathology more broadly.

Our findings also show that, whereas Social withdrawal was positively associated with metacognitive sensitivity, Compulsive hypersensitivity was negatively associated. Reduced metacognitive sensitivity in the transdiagnostic compulsive dimension is consistent with previous clinical and transdiagnostic studies (Seow and Gillan, 2020; Vaghi et al., 2017). Interestingly, the positive association between Social withdrawal and metacognitive sensitivity may indicate heightened monitoring of internal performance signals, analogous to the increased insight into performance fluctuations reported for a depression-anxious dimension (Rouault et al., 2018), rather than reflecting impaired metacognitive processes.

The advantage of examining psychopathology at the transdiagnostic rather than symptom level was particularly evident for metacognitive sensitivity. Symptom-level associations were inconsistent in both magnitude and direction across symptoms loading on the same factor, yet coherent and significant associations emerged at the dimension level, suggesting that transdiagnostic factors capture cognitive profiles that individual symptom scores fail to resolve (Wise et al., 2023). Note, however, that our task involved sequential learning rather than independent trials, which may introduce a systematic shift in the measured metacognitive sensitivity. However, since this effect should be consistent across participants, it is unlikely to bias associations with psychiatric dimensions, although it may attenuate their magnitude. Importantly, significant effects were present even with strict screening of inattentive participants and controlling for response bias, suggesting the findings are robust to general confounding variables (Sarna et al., 2026)

The convergence of diminished abstraction use and reduced metacognitive sensitivity within the Compulsive hypersensitivity dimension points to dysfunction at two distinct but complementary levels of cognitive organisation, each of which represents a potential therapeutic target. At the representational level, the abstraction deficit identified here suggests that recalibrating how task-relevant structure is extracted and encoded may be a promising intervention target. Decoded neurofeedback (DecNef) has been proposed as one useful approach to this end in compulsive disorders (Zhang et al., 2026), operating by reinforcing distributed multivoxel neural representations rather than coarse regional signals (Taschereau-Dumouchel et al., 2022, 2021). Critically, neurofeedback has been shown to directly enhance abstraction by reinforcing neural representations of task-relevant features and increasing their utilisation for decision making (Cortese et al., 2021; Kahnt, 2022), providing a mechanistic rationale for applying DecNef to counteract the feature-level over-reliance characteristic of compulsive psychopathology. At the metacognitive level, interventions targeting maladaptive biases about one’s own thoughts and performance have shown promise for compulsive psychopathology (Seow et al., 2025b). Together, these approaches may be particularly suited to transdiagnostic dimensions in which both representational and metacognitive processes are disrupted (Seow et al., 2025b). Moreover, our computational approach did not incorporate models that integrate abstract and metacognitive representations; future studies should examine whether interactive models (Cortese, 2022) better capture the mechanisms underlying metacognitive dysfunction in psychopathology.

There are some limitations to consider. First, online recruitment may have introduced sampling bias, as participants were limited to those with internet access or an interest in online research. Second, the analytic sample comprised approximately 50% of those initially recruited after applying pre-registered exclusion criteria. However, note that these exclusions followed pre-registered criteria for data quality and should have reduced measurement error (Brühlmann et al., 2020). Third, choice and confidence were reported via the same slider, potentially confounding metacognitive sensitivity, although the absence of a strong initial position effect suggests this is unlikely to account for the observed associations. Fourth, transdiagnostic scores were derived using first-order factor analysis rather than alternative approaches such as bi-factor modelling (Caspi and Moffitt, 2018); nonetheless, first-order factors have been shown to robustly capture cognitive correlates relative to alternative solutions (Fox et al., 2025).

In summary, in this study, we found that a Compulsive hypersensitivity transdiagnostic dimension was characterised by convergent impairments in both abstract task representation and metacognitive evaluation, whereas Social withdrawal was selectively associated with heightened metacognitive sensitivity. These findings support a hierarchical view of cognition in which transdiagnostic psychopathology is also expressed at the level of how task structure is represented and monitored. The dimension-specificity of these associations suggests that representational and metacognitive impairments constitute markers of compulsive psychopathology in particular, with implications for interventions targeting both levels of cognitive organisation.

## Methods

### Transparency, openness, and ethics

The current study has been pre-registered (Open Science Framework repository (OSF), DOI: https://osf.io/qa4bk/). See the supplementary methods for a summary of deviations from the original preregistration. The study has been approved by the Ethics Committee of the Advanced Telecommunication Research Institute (No. 773). Participants provided their informed consent online by clicking “Yes” on the statement “I consent to take part in this study” after reading the plain language statement and consent form.

### Sample size

We used G*power 3.1.9.7 (Franz Faul, Kiel University, Germany) for the a priori power calculator. Because correlations between individual differences in transdiagnostic variables and such RL parameters (*r* = 0.1-0.2) in a similar previous study (Dubois and Hauser, 2022) and the correlation between the transdiagnostic factors and the degree of abstraction RL parameter in our pilot sample (*r = ∼*0.15, N = 15) were low, we assume a one-tailed alpha-level of 0.05, a power of 90%, and a correlation effect size of *r* = 0.15. As a result, the analysis yielded a target sample size of 374, which we rounded to 380. Our estimated number of participants is based on previous online studies using the same approach (Hoven et al., 2023; Seow and Gillan, 2020). At least 25% of subjects in online studies are commonly excluded due to pre-defined exclusion criteria. Moreover, given the unpredictable exclusion rate (∼5%), we recruited 512 subjects. After excluding participants, we obtained 249 participants for our main analysis (see the section on Participant Exclusion Criteria for details). We conducted a post hoc power analysis, and the statistical power was 77%.

### General procedures

To answer our research questions, our experiment assessed a cognitive task and multiple questionnaires in a large-scale, general-population sample recruited online. We recruited the participants online through Prolific (https://www.prolific.co/). The inclusion criteria were that all participants must be fluent in English and aged between 20 and 60 years (note that we preregistered this as 18-65 years). Using Prolific’s system, we only invited participants who reported being fluent in English. Subjects were reimbursed for their participation at an hourly rate and received a bonus based on their performance (see the “Task” section for details). We limited participants’ devices to “Computer” for the task, which can also be set via the Prolific function. We separated the sample into three groups to consider the confounding effect due to time-relevant issues (e.g., chronotype (Mehrhof and Nord, 2024)). Our preregistration stated that the experiment would be conducted in two separate sessions, one in the am and one in the pm, based on UK time. However, to ensure data quality and operational stability, we allocated 250 participants each to the 9:00 AM and 5:00 PM sessions on the same day (June 20, 2025). The remaining 50 participants completed the study at 1:00 PM on the following day (June 21, 2025). This scheduling arrangement was implemented to manage the number of simultaneous participants per session, thereby minimising the potential impact of server load and concurrent access issues. This adjustment does not compromise the validity of comparisons in our main analyses.

### Measures

#### Demographics and questionnaires

We asked about age, sex, marital status, and educational level as demographic information. We also used the following questionnaires:

-Attention/hyperactivity disorder based on Adult ADHD Self Report Scale (ASRS, 18 items) (Kessler et al., 2005)

-Fear/avoid social anxiety symptoms based on Liebowitz Social Anxiety Scale (LSAS-J, 48 items) (Baker et al., 2002)

-Impulsivity based on Barratt Impulsiveness Scale (BIS, 30 items) (Patton et al., 1995)

-Obsessive-compulsive disorder based on Obsessive-Compulsive Inventory (OCI, 42 items) (Foa et al., 1998)

-General mental health (anxiety/depression) based on Kessler Psychological Distress Scale (K6, 6 items) (Prochaska et al., 2012)

-Alcohol and tobacco use disorder based on CAGE (4 items) (Ewing, 1984)

-Tobacco use disorder based on Tobacco Dependence Screener (TDS, 10 items) (Kawakami et al., 1999)

-Gaming disorder based on Problematic Online Gaming Questionnaire (POGQ, 18 items) (Demetrovics et al., 2012)

We have selected these questionnaires, whose scores are relevant to behavioural regulation in general and are closely correlated, given their comorbidity and interrelationships (Oka et al., 2025).

#### Attention check

We also added an attentional check questionnaire to exclude participants who may be inappropriate. The questionnaire is located in the fourth order to check their attention at the intermediate timing. We used only one attentional check questionnaire to minimise unnecessary confounding effects on subsequent questionnaires and tasks (Hauser and Schwarz, 2015).

#### Items to exclude inattentive participants and control biases

Following previous research (Sarna et al., 2026; Zorowitz et al., 2023), we asked several questions to assess participants’ inattentiveness and response biases. For this study, we have already modified the sentences in the original questions to fit into each questionnaire. See supplementary materials for the details.

### Task

#### Abstraction reinforcement learning task

The abstraction task was implemented in Gorilla (Anwyl-Irvine et al., 2020). The task consists of learning the fruit preference of PAC-MAN-like characters. These characters have three features, each with two levels: colour (blue vs yellow), stripe orientation (horizontal vs vertical), and mouth direction (left vs right). On each trial, a PAC-MAN-like character composed of a unique combination of the three features was shown. The experimental session was divided into blocks, each with a specific rule that directed the association between features and preferred fruits. There were six blocks at most. There were two relevant features and one irrelevant feature, but these changed randomly at the beginning of each block. Therefore, overall, there were three types of blocks (each classified using its two relevant features): CD (colour-direction), CM (colour-mouth direction), and DM (direction-mouth orientation). For example, ‘blue + vertical stripes → grapes’, in which case the mouth orientation is not related to the food preference. Each rule appeared twice in each of the six blocks, and the order was randomised (see Supplementary Figure 1 for more detail). Furthermore, to avoid a simple logical deduction of the rule after one trial, the four possible combinations of two relevant features with two levels were paired with the two fruits in both a symmetric and asymmetric fashion, i.e., 2×2 or 3×1 (see Supplementary Figure 1 for the visualised tables). Moreover, the stimuli of the two fruits were also randomly changed at the beginning of each new block.

Each trial starts with a fixation screen for 2s, followed by the Pacman-like character being presented for 1s. Then, while the character remains at the centre of the screen, the two fruit options appear below it on the right and left. Participants had 5s to move a cursor to their selected position on a horizontal bar (see Supplementary Figure 2 for the average of response duration). The selected position indicates the participant’s inference of the character’s preference (right for the fruit to the right, left for the fruit to the left), as well as the participant’s confidence in this inference (more central position = lower confidence, more lateral position = higher confidence, see Supplementary Figure 3 for the distribution).

Finally, the feedback was provided, and a tick appears on the screen if the selection is correct; otherwise, a cross appears. The feedback remains on screen for 1s, following which the trial ends with a variable ITI, bringing the trial to a fixed 8s duration.

Participants started with 40 practice trials. While each block contains up to 60 trials, a block can end earlier if participants learn the target rule. Learning is defined as a consecutive set of correct trials (ranging from 8 to 12, determined randomly in each block). Participants were instructed to receive one point for each correct choice and zero points (i.e., no reward and no penalty) for an incorrect choice. The session ended after 30 minutes, even if participants did not finish all blocks. They were further informed that bonus points were assigned, weighted by learning speed within each block. That is, the faster the rule is learned, the more points are awarded. Participants were compensated with a base payment of £6 plus a bonus (£4.2 at most) based on their performance in the abstraction task. The bonus reward was computed at the end of the session according to the formula:

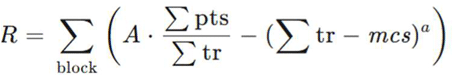

*R* is the reward obtained in that block. A is the maximum available reward in a block (£0.7). ∑pts represents the sum of correct responses, and ∑tr represents the number of trials participants use in the block. *mcs* is the maximum length of a correct strike (12 trials), and *a* is a scaling factor (*a* = 0.005). This formula ensures time-dependent decay of the reward, which approximately follows a quadratic fit. If participants completed a block in fewer than 10 trials, the amount that would otherwise have been larger than £0.7 was rounded to £0.7. If participants do not achieve five consecutive correct answers in the practice session, they can attempt one more practice block.

We used the comprehension questions after the practice to confirm the understanding of the confidence rating. To fit into our task style, we have modified the questions that previous studies have used:

1. If you are certain you made the correct judgment, where on the scale would you place your confidence, from around the centre (‘Guessing’) to either edge (‘Certainly correct’)?
2. If you are completely unsure whether you made a correct judgment, where on the scale would you place your confidence, from around the centre (‘Guessing’) to either edge (‘Certainly correct’)? Participants must answer both questions correctly, not just one.

### Controlling for cognitive confounds

We have considered including IQ as a covariate in our analyses, given previous studies reporting associations between individual differences in model-based behaviour and general intelligence (Gillan et al., 2016). However, in the context of the present study, our primary dependent variable, the abstraction-related parameter, functionally overlaps with specific components of IQ. Controlling for IQ in this context could therefore lead to overadjustment bias, in which meaningful variance related to the cognitive processes of interest (i.e., abstraction in decision-making) is inadvertently removed. Instead, we included the Deary–Liewald Reaction Time (DLRT) task (see Supplementary methods for the details) as a covariate to account for basic processing speed, which may influence performance on our task but does not overlap with the higher-order abstraction processes central to our hypotheses. This approach allows us to control for lower-level confounds while preserving the variance in the cognitive processes we aim to investigate.

### Participant Exclusion Criteria

Participants were excluded from the primary analyses based on the following criteria:

1. Exclude participants whose average performance is at or below chance (as measured by a binomial test within participants): 68
2. Exclude participants with an average reading time of the primary instruction page of <5 seconds: 25
3. Double attention check failure (Attention check + infrequency item mistake more than 2): 203
4. Participants who responded with the same value (e.g., all 1s or all 4s) to all items in a questionnaire that included reverse-coded items were excluded from analysis based on inattentive responding: 0
5. Exclude participants who fail to complete the blocks within the allocated time (30 minutes): 0
6. Exclude participants who fail comprehension questions on the confidence scale: 58
7. Exclude participants who always choose the same location 80% of the time: 0

We did not use the DLRT results for this exclusion because of many unexpected participant errors (see below), which deviated from our preregistration.

### Analysis

#### Computational modeling

We used Hierarchical Bayesian Inference (HBI) to fit the models to individual behavioural data, enabling us to estimate hyperparameters block-wise precisely (Piray et al., 2019). Following Cortese et al. (2021), we construct a classical reinforcement learning (RL) model called Q-learning. Here, a state is defined as a combination of features. The feature RL model has 2^3^ = eight states, where each state is given by the combination of all three features. The abstract RL model is designed with 2^2^ = 4 states, where each state is defined by the combination of two features. These learning models create individual estimates of how participant action selection depended on features they attended and their past reward history. Both RL models share the same underlying structure and are formally described as:

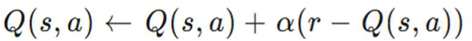

where *Q(s,a)* is the value function of selecting either fruit-option *a* for PAC-MAN state *s*. The value of the action selected in the current trial is updated based on the difference between the expected value and the actual outcome (reward or no reward), known as the reward prediction error (RPE). Expected values of the two actions are combined to compute the probability *P* of predicting each outcome using a SoftMax (logistic) choice rule:

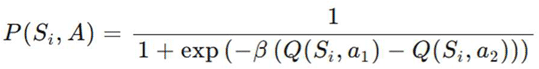

The greediness hyperparameter *β* controls how much the difference between the two expected values actually influences choices. That is, hyperparameters estimated through likelihood maximisation were the learning rate *α* and the greediness (inverse temperature) *β*. Abstraction ability was calculated as the average ratio of the best model fitting across blocks at each participant level. To validate the model parameters, we conducted a parameter recovery analysis following established procedures (Palminteri et al., 2017). Choice data were simulated for each participant and block using their best-fitting parameters estimated from the main analysis. To mimic trial-by-trial variability, noise was introduced during simulation, with Q-value updates occasionally applied to a randomly selected state rather than the true task state (noise probability = 0.1). Simulated data were then refitted using the same HBI procedure as in the main analysis, separately for each model. Recovered and original parameter values were pooled across participants, blocks, and models, and Spearman correlations between original and recovered parameters were computed to assess recovery quality. Our pre-registration specified a sensitivity analysis that included excluded participants, and these participants differed significantly on several psychiatric scores (e.g., ADHD, impulsivity; p < 0.001). Nevertheless, this analysis was not conducted because the hierarchical Bayesian model pools information across participants, meaning that including them would alter parameter estimates for the retained sample. We therefore focus on the pre-defined, strictly screened sample.

#### Statistical analysis

We conducted three bootstrap multiple linear regressions (5,000 iterations, case-resampling with replacement, preserving the joint covariate distribution) based on a generalised linear model to examine the relationship between task-related measures as dependent variables (parameters) and 1) psychiatric symptoms and 2) transdiagnostic dimensional scores. The entire regression model was re-estimated at each iteration, and two-tailed bootstrap *p*-values were computed as 2 × min (*p_left*, *p_right*), where *p_left* and *p_right* denote the proportions of bootstrap samples falling below and above zero, respectively. In these regression models, we also controlled for age, sex, educational level (treated as an ordinal variable), and processing speed (calculated from phase 1 of the DLRT task) as fixed effects. Note that we did not use Phase 2 data in the DLRT task because numerous participants had inadvertently skipped the instruction, and we excluded a random effect for participants from our pre-registered models because the term was meaningless for these between-participant analyses. We also applied a multiple-comparison adjustment based on the false discovery rate (Benjamini and Hochberg, 1995) to symptom-based analyses, as we repeated the model testing across scores.

Specifically, the model equations were defined as follows (Wilkinson notation),

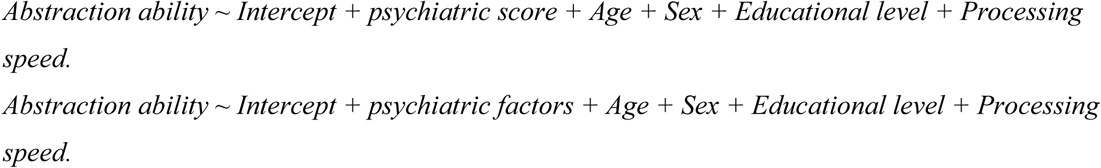

To avoid multicollinearity, the independent psychiatric score was input separately, e.g.:

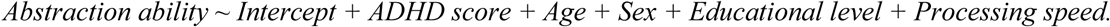

Psychiatric factor scores were input together, i.e., Factor 1 + Factor 2 + Factor 3.

We explored the relationship between psychiatric scores/factors and confidence (i.e., based on the distance from the cursor location to the centre of the slider). The confidence level was calculated as the average across blocks. We added the averaged response to content-neutral items as a control variable to account for general response tendencies because recent research suggested that failing to control for such biases might distort the association between confidence and subjective psychopathology (Sarna et al., 2026).

Specifically, the model equations are defined as follows,

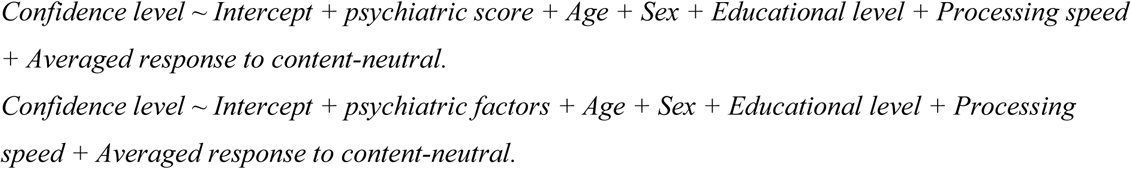

Additionally, we calculated metacognitive sensitivity based on confidence. We applied the area under the type 2 ROC curve (AUROC2) as a metacognitive sensitivity metric and input it as the dependent variable. Note that participants (N = 5) who used only a single confidence rating across all trials were excluded from AUROC2 computation, as the Type-2 ROC curve cannot be defined in the absence of rating variance. These participants were, however, retained in analyses using mean confidence ratings, as invariant responding may itself reflect a meaningful metacognitive response style.

Specifically, the model equations are defined as follows,

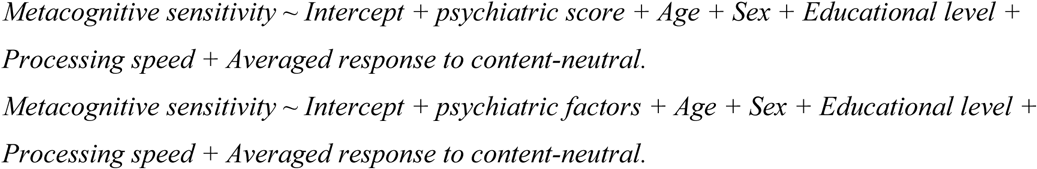

Moreover, to explore the interaction effects between metacognitive sensitivity and psychiatric scores for abstraction ability. These models did not improve the model fit according to the likelihood ratio test (see Results and Supplementary Table 7).

## Author details

**Taiki Oka**

Contribution: Conceptualisation, Resources, Data curation, Software, Formal analysis, Funding acquisition, Validation, Investigation, Visualisation, Methodology, Writing - original draft, Project administration, Writing - review and editing

For correspondence

Conflicts of interest:

There are no conflicts of interest.

**Yoshihiko Kunisato**

Contribution:

Conceptualisation, Software, Formal analysis, Supervision, Validation, Methodology, Writing - review and editing

Conflicts of interest:

There are no conflicts of interest.

**Kotaro Koizumi**

Contribution: Formal analysis, Validation, Visualisation, Writing - review and editing Conflicts of interest: There are no conflicts of interest.

**Misa Murakami**

Contribution:

Resources, Investigation, Writing - review and editing Conflicts of interest: There are no conflicts of interest.

**Hugo Six**

Contribution:

Methodology, Writing - review and editing Conflicts of interest: There are no conflicts of interest.

**Jessica E Taylor**

Contribution:

Formal analysis, Investigation, Methodology, Writing - review and editing Conflicts of interest: There are no conflicts of interest.

**Aurelio Cortese**

Contribution:

Conceptualisation, Software, Formal analysis, Supervision, Funding acquisition, Validation, Methodology, Writing - original draft, Project administration, Writing - review and editing

For correspondence

Conflicts of interest:

There are no conflicts of interest.

## Data and code availability statement

Data, relevant scripts for all analyses, and other supplementary materials can also be found on OSF (DOI: osf.io/qa4bk/).

## Acknowledgement

Funders played no role in the study design, data collection, analysis, or interpretation. We thank Miho Nagata for their help scheduling the experiments. We also thank Mor Ovadia for helping with the implementation of experiments.

