## Supplementary materials for "Mapping abstraction and metacognition onto distinct transdiagnostic symptom profiles"

#### Supplementary methods

##### Participants characteristics

The final sample's characteristics are as follows:
The gender distribution was male 134 (53.8%), female 106 (42.6%), and non-respondents 9 (3.6%). The average age was 32.9 (SD: 9.8). The distribution of their countries of residence was as follows: South Africa: 34.4%; United States: 22.8%; United Kingdom: 17.4%; Poland: 5.4%; and others: 22.5%. Please note that we were unable to obtain demographic information from 9 people.

##### Measurement

**Infrequency items**

1. I was worried about the leprechauns who guard the hidden treasure (expected answer: 'not at all', which was included in OCI (35th question).
2. How often do you rearrange the furniture in my home to prepare for the arrival of magical beans? (expected answer: 'Never'), which was included in ASRS (12th question).
3. I am worried about the canine World Cup (expected answer: 'Rarely/Never'), which was included in BIS (31st question).
4. How much time did you spend worrying about the 1977 Olympics over the last two weeks? (expected answer: Never), which was included in POGQ (8th question).

Given the structure of each questionnaire (e.g., the small number of items), we did not include such items in other questionnaires (LSAS, K6, CAGE, and TDS) because the insertion of an infrequent item might affect their reliability. We did not reward participants who failed the above attention check and the infrequency question in BIS and asked them to return their submission, following Prolific's attention and comprehension check policy (Gagné and Franzen, 2023; Team, 2022).

**Content-neutral items**

Based on previous research(Sarna et al., 2026), we also used content-neutral items to assess participants' acquiescence tendencies, ranging from 1 ("Strongly Disagree") to 5 ("Strongly Agree"). We have excluded one question ('As I see it, Madrid is a small place.') from the original question sets because it requires prior knowledge (i.e., participants must know what "Madrid" is to answer this question). See Supplementary materials for the details.

1. I think quantitative information is difficult to understand.
2. When I go shopping, I find myself spending very little time checking out new products and brands.
3. Everyone should use a mouthwash to help control bad breath.
4. A college education is very important for success in today's world.
5. I like to visit places that are totally different from my home.

6. I work very hard most of the time.
7. I will probably have more money to spend next year than I have now.
8. I think fashion is irrelevant.
9. I believe there are relatively few different breeds of cats.
10. I think the moon is very far from Earth.
11. Book covers are important, in my opinion.
12. These days, matchboxes are no longer useful.
13. I find the taste of apples different from that of pears.

We randomised the order of questionnaires across participants to avoid order confounding effects, except for the attentional check.

##### Abstraction reinforcement learning task

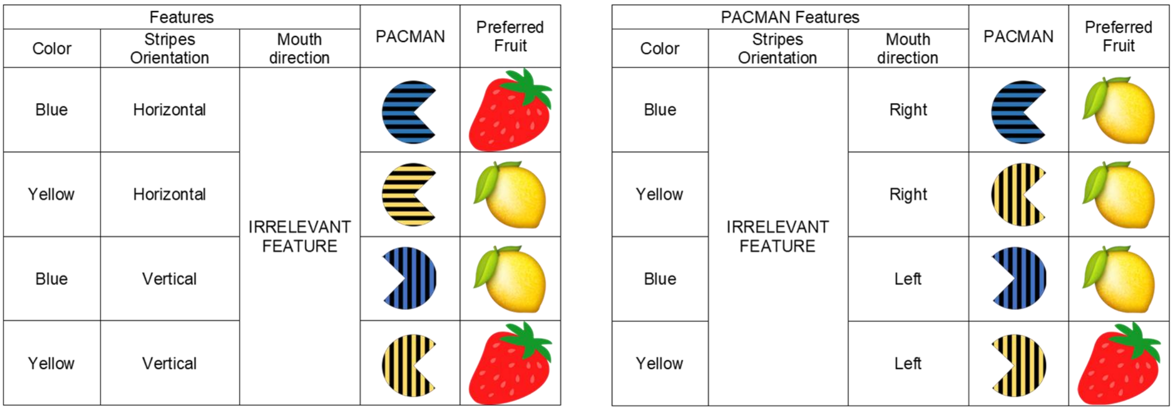


**Supplementary Figure 1 Feature combinations for each condition (2x2 or 3x1)**

*To avoid a simple logical deduction of the rule after one trial, the four possible combinations of two relevant features with two levels were paired with the two fruits in both a symmetric and asymmetric fashion, i.e., 2x2 or 3x1*

##### Control task: Deary-Liewald reaction time task (DLRT)

To control simple or choice processing speed, we conducted the Deary-Liewald reaction time task (DLRT)(Deary et al., 2011). In the first task, participants must press the “spacebar” key upon stimulus appearance (a cross in a square on the computer screen). After pressing the key, the cross in the square disappears and reappears after a short interval. The stimulus interval is randomly set to a value between 0.4 and 2 seconds. In this task, 8 practice trials and 20 main trials were performed, following previous studies (Ferreira et al., 2021). The median RT(ms) of each participant was used in the analysis as a measure of simple processing speed. In the second task, four white squares were centred and aligned horizontally on a computer screen. The leftmost square corresponds to the "z" key, the second square from the left to the "x" key, the second square from the right to the "comma" key, and the rightmost square to the "full-stop" key. On each trial, a cross appears at random in one of the squares, and the participant must press the corresponding key as quickly as possible. When the participant presses one of the four keys, the cross disappears, and another appears on the screen after a short delay. The stimulus interval is randomly set to a value between 0.4 and 2 seconds. The task includes eight practice trials and 40 test trials. The median RT (ms) was used as a measure of choice processing speed in the data analysis. This task result was used to control processing speed and to exclude participants who exhibit problematic behaviours, such as answering randomly or cheating with bots (e.g., generative AI; see Exclusion Criteria in the main text). Regarding the second task, reports indicated that buttons were not responding properly or that tasks started without participants being able to read the instructions due to design issues. Since there was an enormous amount of data showing no response, exclusion criteria based on this were not adopted.

#### Statistical analyses

##### Exploratory factor analysis

We applied a de novo exploratory factor analysis with maximum likelihood estimation (MLE) to confirm whether the factor structure is consistent with that in our previous study (Oka et al., 2025), which was based on a large online Japanese population. Although we have already obtained the three factors and their weights using the same questionnaires, almost all participants in this study were not Japanese. We must confirm the consistency (or inconsistency) across languages, countries, cultures, and ethnicities. This analysis was conducted using oblique rotation (OBLIMIN), with the 176 question items being entered as variables. For this factor analysis, we used unscaled data to compute the heterogeneous correlation matrix, since several scales use binary ratings. We used weights derived from larger online samples (Oka et al., 2025) to transform the scores of our experimental sample to obtain transdiagnostic scores. Note that the expression “confirmatory” factor analysis in our preregistration was wrong.

##### Statistical models for behavioural analyses

To analyse the relationship between block number (time) and learning speed, we conducted a linear mixed-effects regression.

The formula is:

$$Learningspeed \sim time + (1 + time \mid participantID)$$

To analyse the relationship between averaged confidence across trials and learning speed, we conducted a robust linear regression. These analyses to check task validity deviated from our preregistration because the original analysis was deemed inappropriate for assessing the task's replicability due to differences in the number of blocks.

##### Deviations from Preregistration

The following deviations from the preregistered protocol (https://osf.io/qa4bk/) were made. Each deviation is described, along with the rationale for the change.

**1. Age range** The preregistered age range was 18–65 years; the analysed sample was restricted to 20–60 years. This restriction reflected the approved age range specified in the ethics documentation, which took precedence over the preregistered range.

**2. Session structure** Two sessions (AM/PM, UK time) were preregistered; data were collected across three sessions over two days. This change was implemented to accommodate participants across multiple time zones, thereby distributing session times and minimising the potential confounding influence of chronotype.

**3. Statistical approach** Standard multiple regression was preregistered; bootstrap regression was used in the main analyses. Bootstrap regression was preferred as it is more conservative and does not assume normality of residuals, providing more robust inference given our sample size.

**4. Exclusion criteria** DLRT-based exclusion was preregistered but was not applied, as a substantial proportion of participants skipped task instructions, rendering the DLRT unusable as a quality metric.

**5. Sensitivity analysis: excluded participants** Our preregistration initially indicated that a sensitivity analysis including excluded participants would be conducted, especially given that the included and excluded samples differed significantly in attentional and impulsivity-related measures (ASRS and BIS, both $p < .001$). However, this analysis was not performed. The hierarchical Bayesian model used here makes such an analysis less informative, because including excluded participants would alter the parameter estimates for all participants, rendering a direct comparison with the main analysis uninformative.

**6. Abstraction ability metric** The preregistered metric was the proportion of blocks in which the Abstract RL model provided a better fit than the Feature RL model. We instead used the mean posterior responsibility of the Abstract RL model across blocks as the primary metric, since it takes only 7 discrete values, limiting sensitivity to individual differences.

### Supplementary results

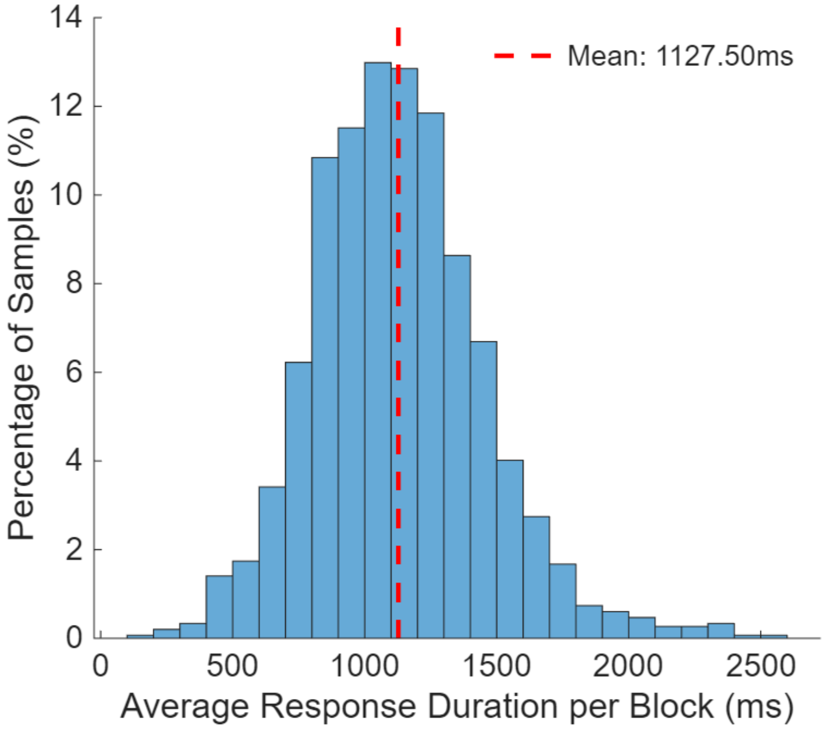


**Supplementary Figure 2. Distribution of response duration across participants**

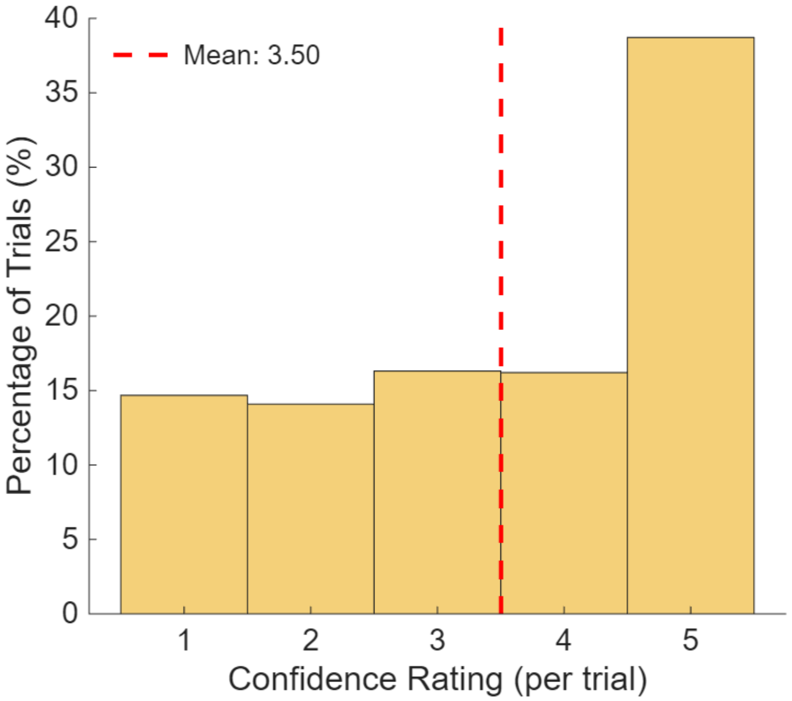


**Supplementary Figure 3. Distribution of confidence ratings**

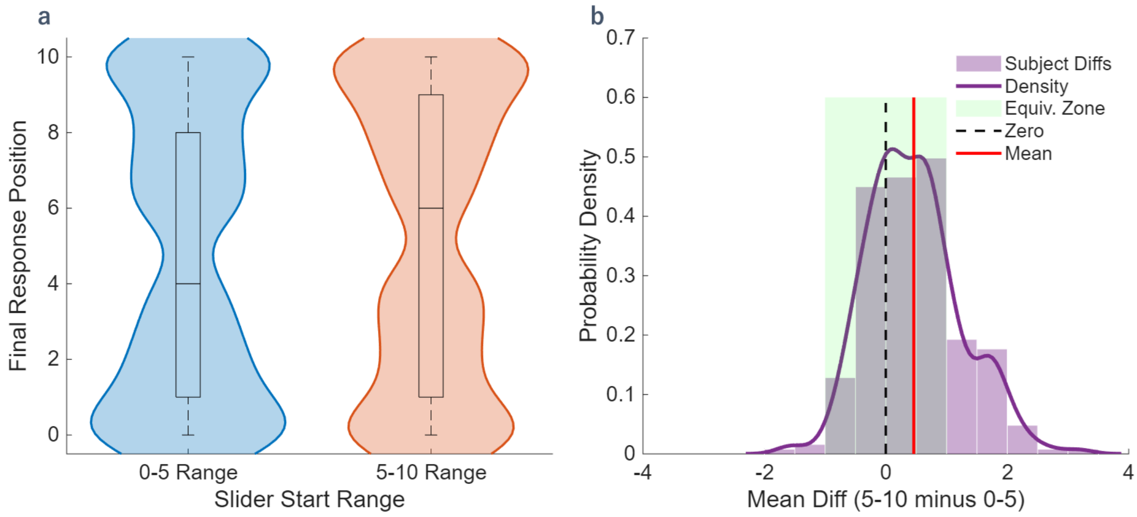


**Supplementary Figure 4. Difference in response by each starting location of the circle on the bar.**

*a. a violin plot between the starting locations b. Distribution of the difference between the initial positions and the average of the final cursor positions. Model (Response ~ Slider_start_min + (1 + Slider_start_min | ParticipantID)) estimates the effects of the starting location. Although the anchoring effect is significant ($\beta$ = 0.093, $p < 0.001$), it is a very small effect size; the variance attributable to the anchoring effect accounts for only 0.03% of the total variance in cursor movement, including the residual variance.*

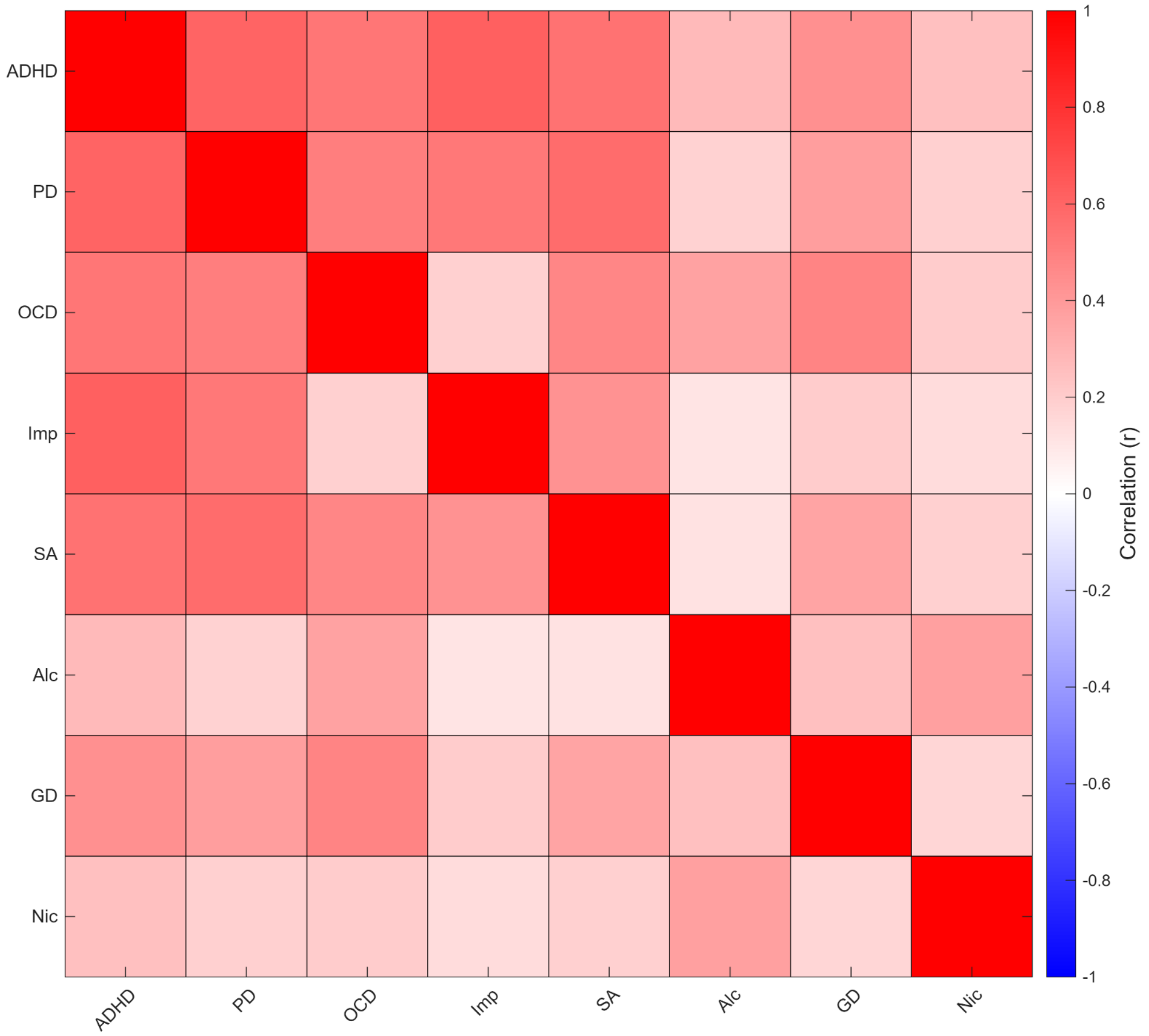


**Supplementary Figure 5. Correlation matrix between psychiatric scores**

*ADHD attentional deficits/hyperactivity disorder; SA social anxiety symptoms; Imp Impulsivity; OCD obsessive-compulsive disorder; PD Psychological distress; Alc alcohol-related disorder; Nic Nicotine dependence; GD Gaming disorder.*

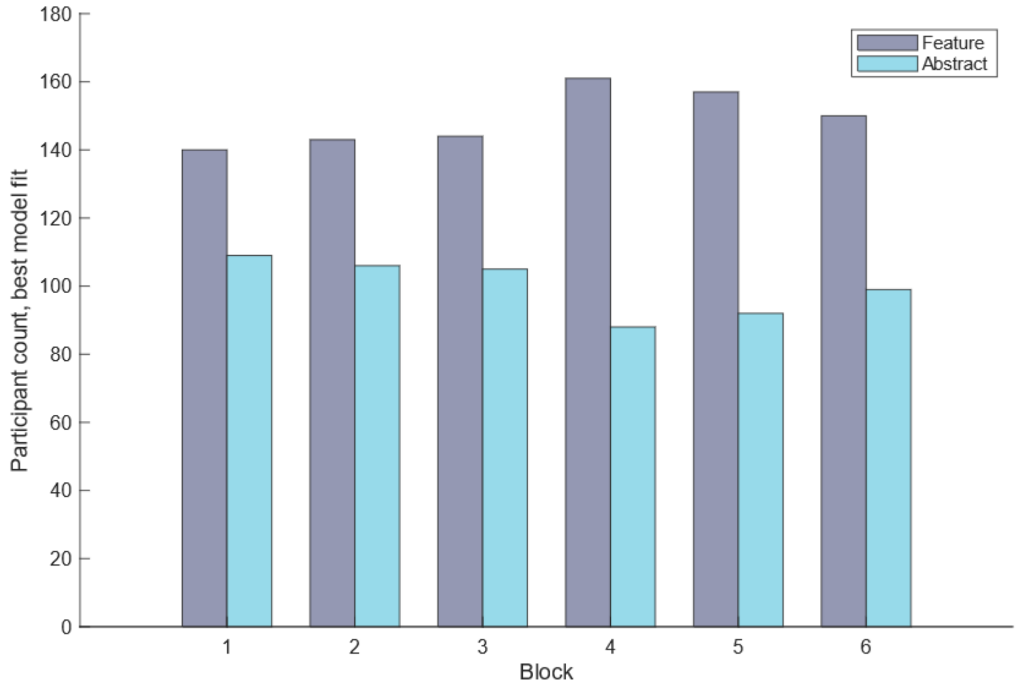


**Supplementary Figure 6. Participants count for the best-fitting model in each block.**
*Participant count for the best-fitting model in each block. The bars show the number of participants for whom the Abstract RL model provided a better fit to behaviour than the Feature RL model in each block.*

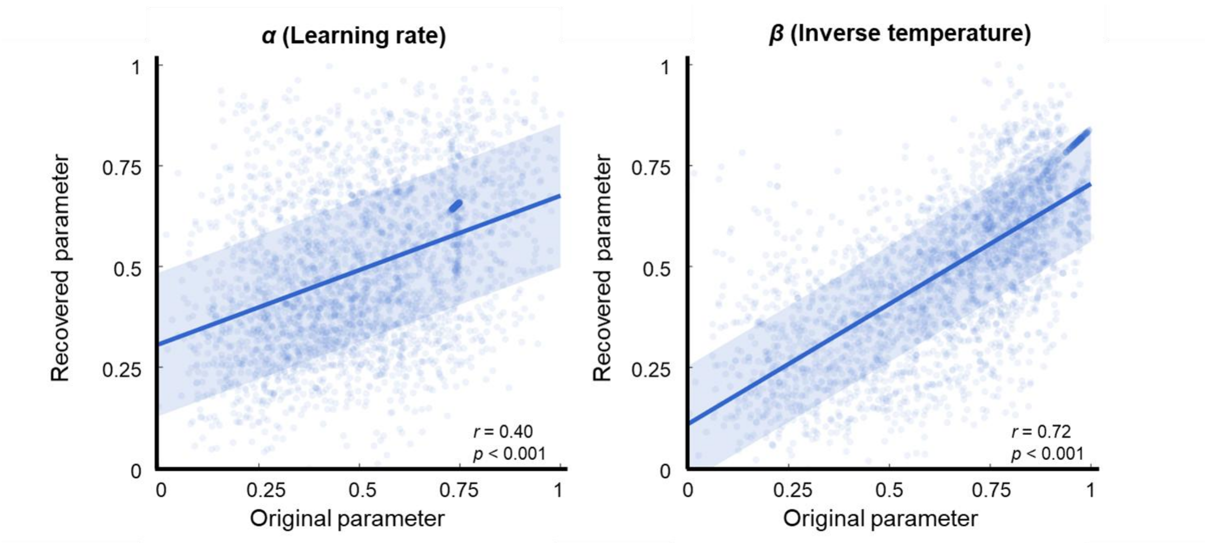


**Supplementary Figure 7 Parameter recovery**. *Choice data were simulated using best-fitting parameters with added noise. Models were then refit to simulated data using HBI separately for each model. Recovered and original parameter values were pooled across participants, blocks, and models, normalised to [0, 1], and plotted against each other. The line represents the linear fit, and the shaded region denotes ±1 SD of residuals. Spearman's correlation coefficients are shown for each parameter.*

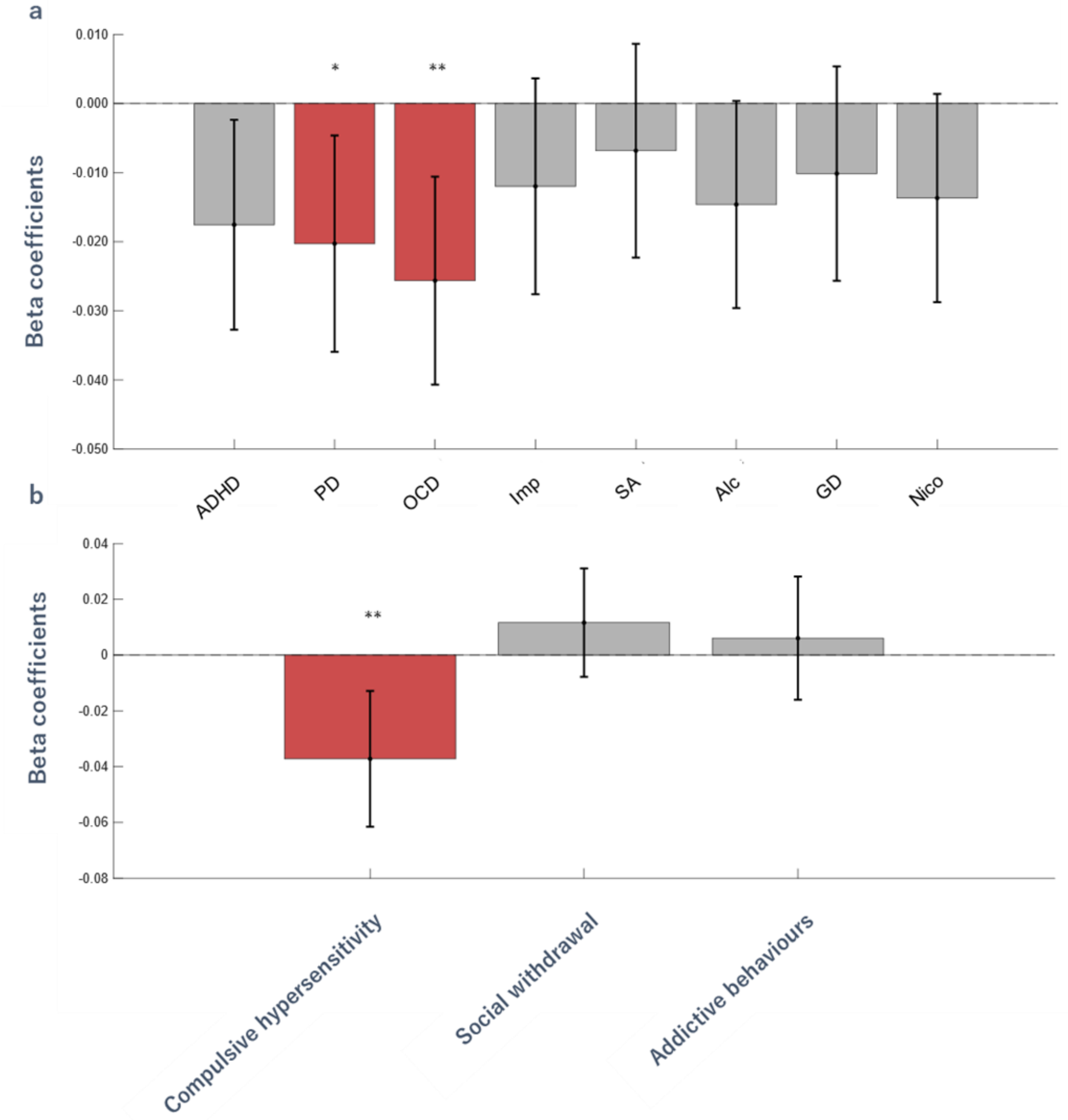


**Supplementary Figure 8. Associations between abstraction ability and psychopathology.**

*Regression coefficients between abstraction and questionnaire scores and dimension scores without the bootstrap method.* ***a. Between the abstraction parameter and each symptom score.*** *ADHD attentional deficits/hyperactivity disorder; PD Psychological distress; OCD obsessive-compulsive disorder; Imp Impulsivity; SA social anxiety symptoms; Alc alcohol-related disorder; GD Gaming disorder; Nic Nicotine dependence.* ***b. Between the abstraction parameter and each dimension score.*** *Error bars denote 95% confidence interval. ♯ represents* $p_{uncorr} < 0.01$.

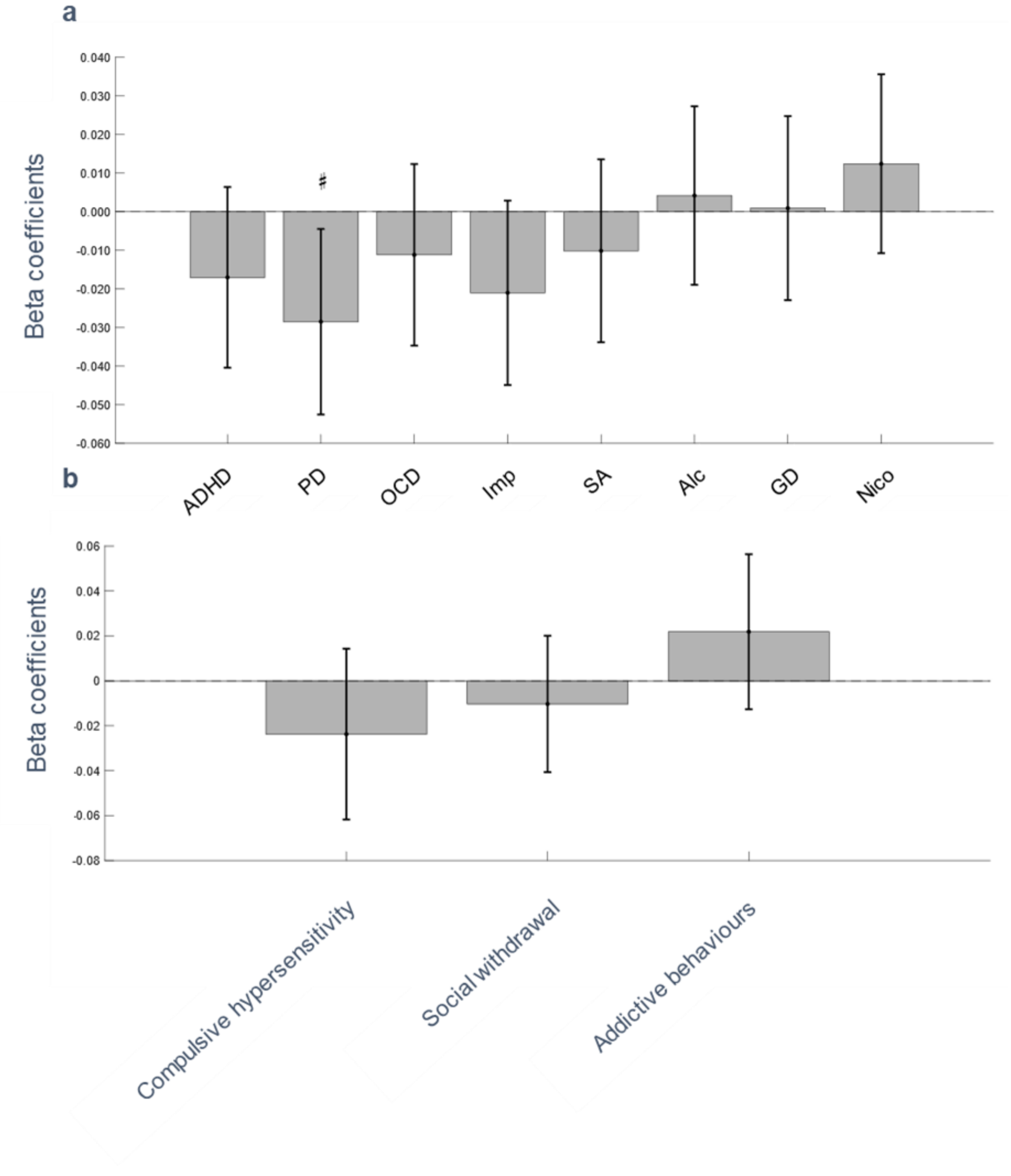


**Supplementary Figure 9. Associations between abstraction ability and psychopathology using the preregistered discrete metric.**

*Regression coefficients between the proportion-based abstraction metric (the proportion of blocks in which the Abstract RL model provided a better fit than the Feature RL model) and questionnaire and dimension scores, reported for transparency as a preregistered sensitivity analysis (without bootstrapping).* ***a. Between the abstraction parameter and each symptom score.*** *ADHD attentional deficits/hyperactivity disorder; PD Psychological distress; OCD obsessive-compulsive disorder; Imp Impulsivity; SA social anxiety symptoms; Alc alcohol-related disorder; GD Gaming disorder; Nic Nicotine dependence.* ***b. Between the abstraction parameter and each dimension score.*** *Error bars denote 95% confidence interval. ♯ represents* $p_{uncorr} < 0.01$.

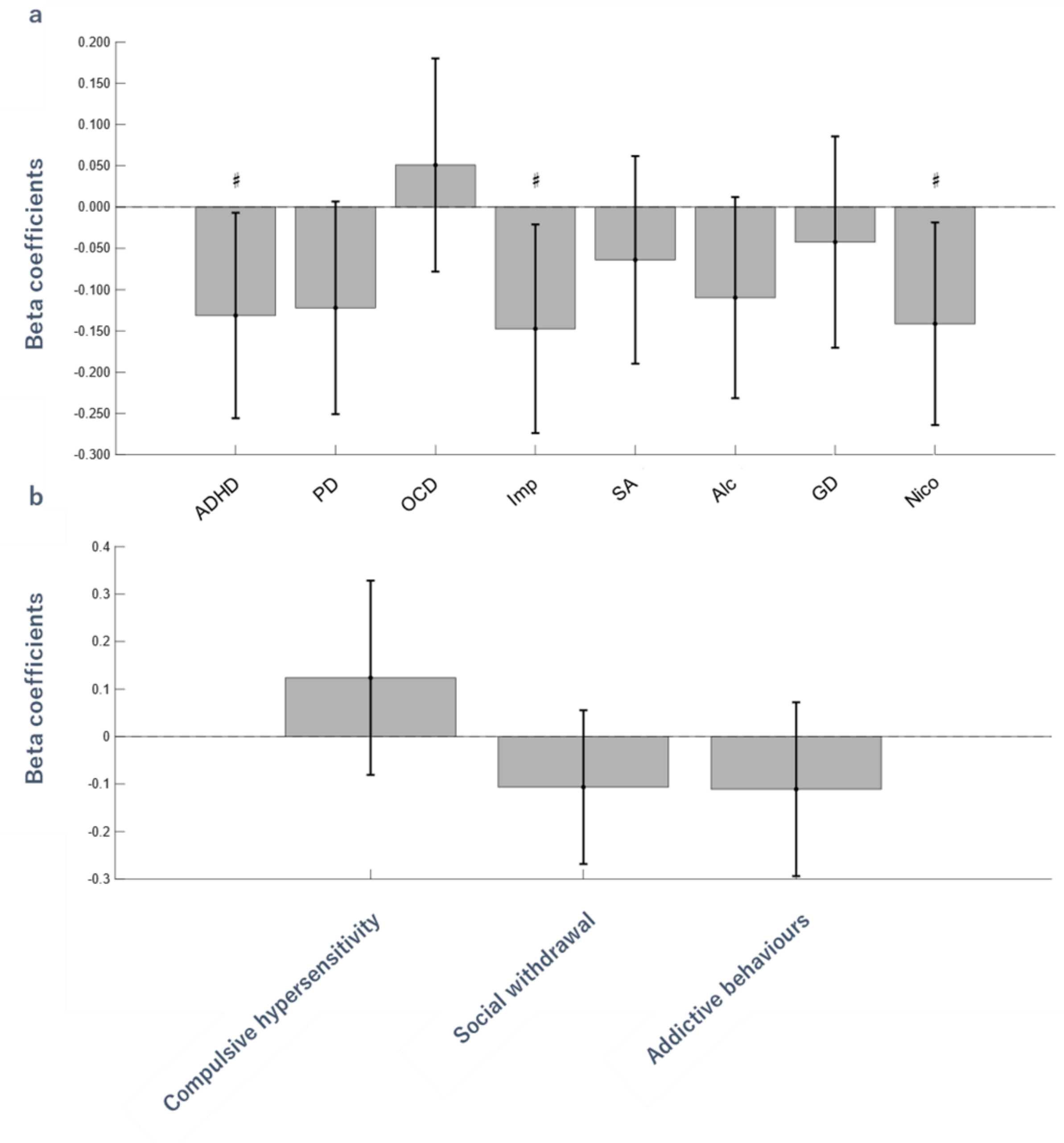


**Supplementary Figure 10. Associations between mean confidence and psychopathology.**

*Regression coefficients between metacognitive bias/sensitivity and questionnaire scores, as well as dimension scores without the bootstrap method.* ***a. Between the averaged confidence and each symptom score.*** *ADHD attentional deficits/hyperactivity disorder; PD Psychological distress; OCD obsessive-compulsive disorder; Imp Impulsivity; SA social anxiety symptoms; Alc alcohol-related disorder; GD Gaming disorder; Nic Nicotine dependence.* ***b. Between the averaged confidence and each dimension score****. Error bars denote 95% confidence interval. ♯ represents* $p_{uncorr}$ *<0.01.*

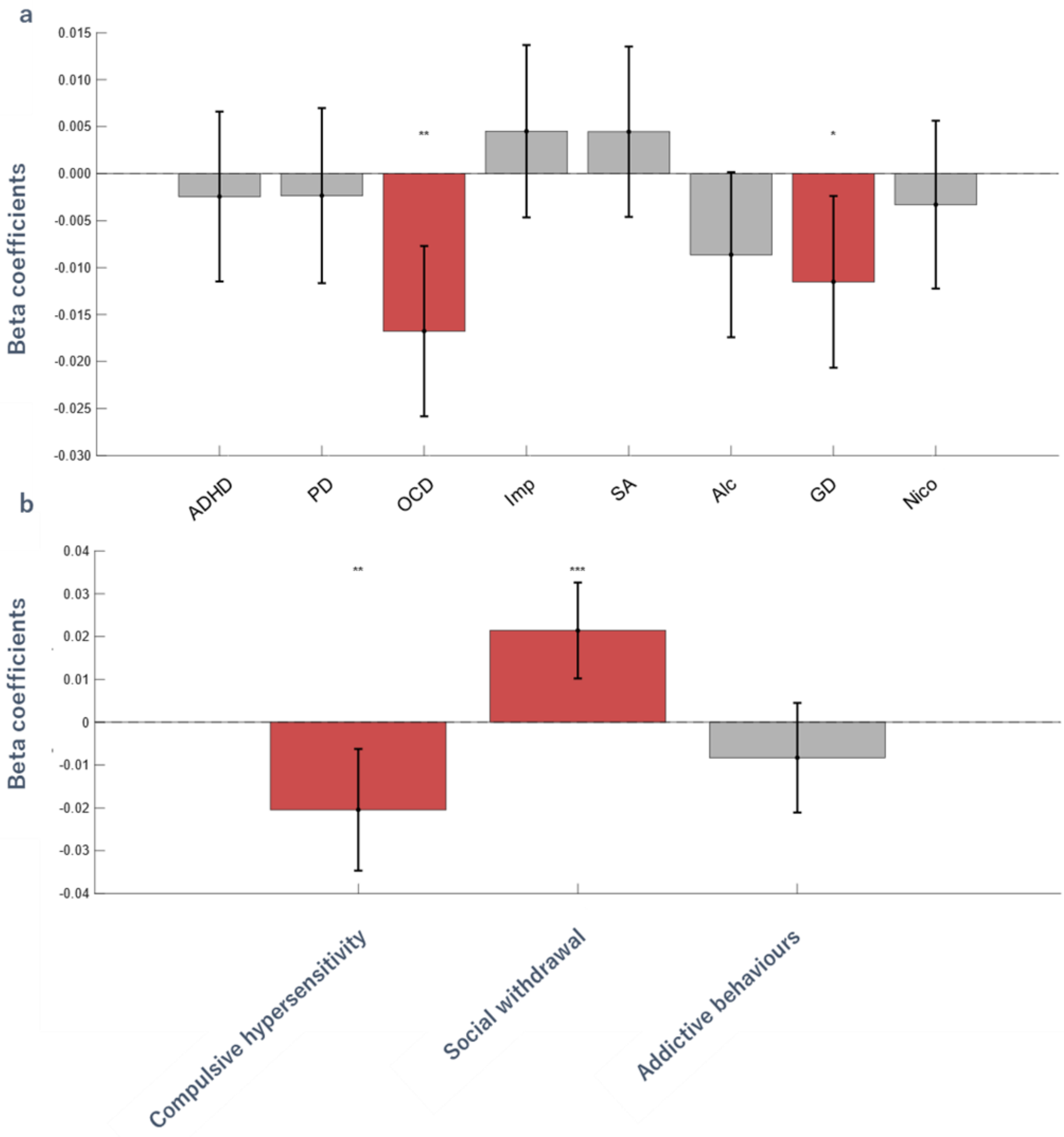


**Supplementary Figure 11. Associations between metacognitive sensitivity and psychopathology.**
*Regression coefficients between metacognitive bias/sensitivity and questionnaire scores, as well as dimension scores without the bootstrap method.* ***a. Between the metacognitive sensitivity and each symptom score.*** *ADHD attentional deficits/hyperactivity disorder; PD Psychological distress; OCD obsessive-compulsive disorder; Imp Impulsivity; SA social anxiety symptoms; Alc alcohol-related disorder; GD Gaming disorder; Nic Nicotine dependence.* ***b. Between the metacognitive sensitivity and each dimension score****. Error bars denote 95% confidence interval. ** $p < 0.01$, *** $p < 0.001$*

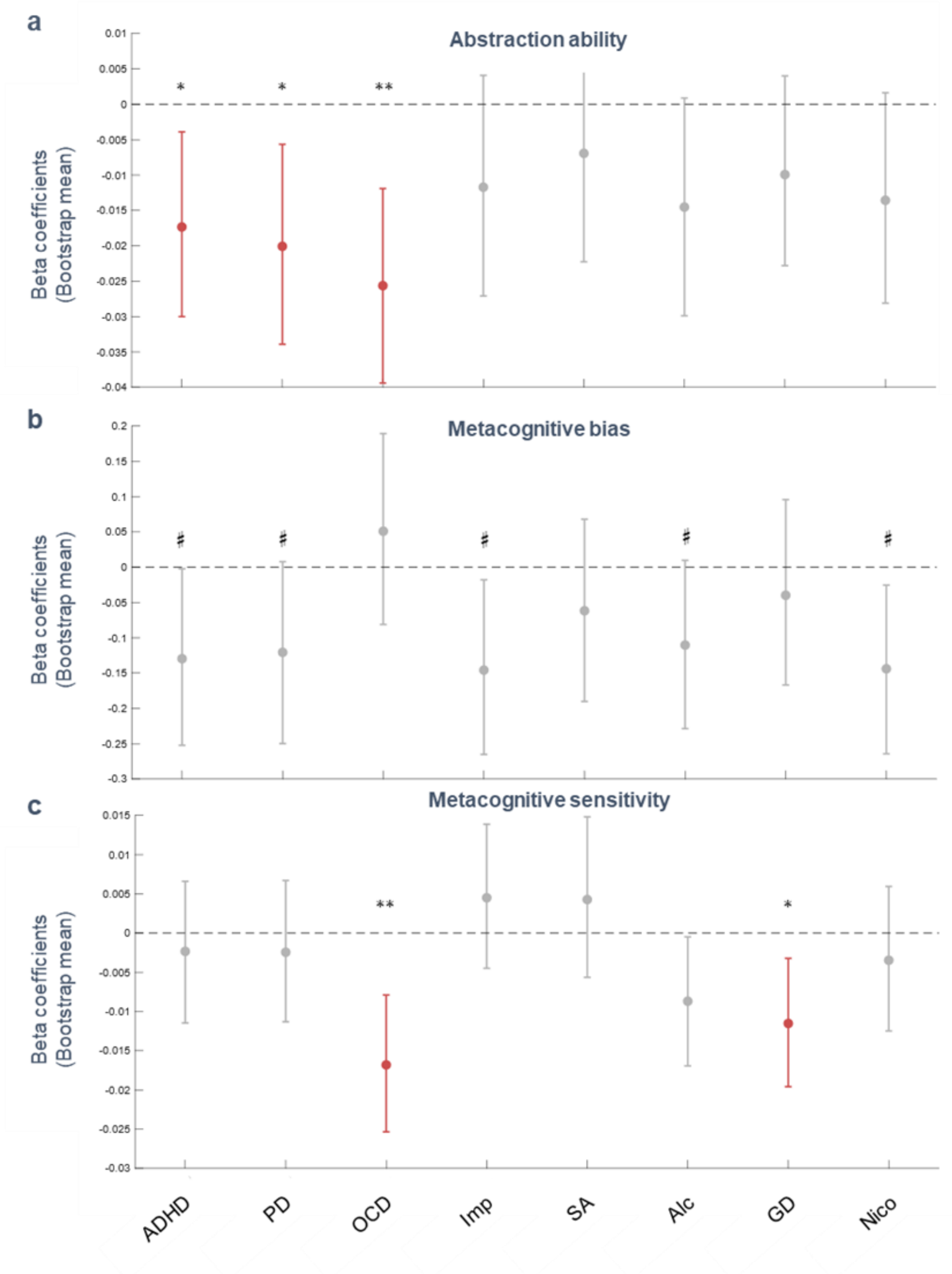


**Supplementary Figure 12 Associations between abstraction ability and symptom-level scores with the bootstrap method.**

*Regression coefficients between each metric and dimension scores based on bootstrap multiple linear regression estimation.* ***a. Between the abstraction parameter and each symptom-level score. b. Between the averaged confidence and each symptom-level score. c. Between the metacognitive sensitivity and each symptom-level score.*** *** $p_{boot} < 0.01$. * $p_{boot} < 0.05$ after multiple comparison adjustments. # represents $p_{uncorr} < 0.01$.*

| | Coefficient | CI lower | CI upper | $p_{boot}$ |
|---|---|---|---|---|
| ADHD | -0.0173 | -0.0300 | -0.0039 | 0.012 |
| Psychological distress | -0.0201 | -0.0339 | -0.0057 | 0.006 |
| Obsessive-compulsive disorder | -0.0256 | -0.0394 | -0.0119 | 0.000 |
| Impulsivity | -0.0117 | -0.0271 | 0.0041 | 0.147 |
| Social anxiety | -0.0069 | -0.0223 | 0.0081 | 0.377 |
| Alcohol-related disorder | -0.0145 | -0.0299 | 0.0009 | 0.065 |
| Gaming disorder | -0.0099 | -0.0228 | 0.0040 | 0.153 |
| Nicotine-related disorder | -0.0136 | -0.0281 | 0.0016 | 0.079 |
| Compulsive hypersensitivity | -0.0372 | -0.0617 | -0.0118 | 0.003 |
| Social withdrawal | 0.0119 | -0.0077 | 0.0314 | 0.230 |
| Addictive behaviours | 0.0061 | -0.0150 | 0.0279 | 0.592 |

**Supplementary Table 1 Statistical values of the associations between abstraction ability and psychopathology.**

*Based on the results without the bootstrap method. Note that the p-value is not adjusted for the purpose of the reference.*

| | Coefficient | CI lower | CI upper | *p*-value |
|---|---|---|---|---|
| ADHD | -0.0170 | -0.0404 | 0.0064 | 0.154 |
| Psychological distress | -0.0285 | -0.0525 | -0.0045 | 0.020 |
| Obsessive-compulsive disorder | -0.0112 | -0.0347 | 0.0124 | 0.351 |
| Impulsivity | -0.0210 | -0.0449 | 0.0029 | 0.085 |
| Social anxiety | -0.0101 | -0.0338 | 0.0136 | 0.401 |
| Alcohol-related disorder | 0.0042 | -0.0189 | 0.0273 | 0.721 |
| Gaming disorder | 0.0009 | -0.0229 | 0.0248 | 0.938 |
| Nicotine-related disorder | 0.0124 | -0.0107 | 0.0356 | 0.291 |
| Compulsive hypersensitivity | -0.0237 | -0.0617 | 0.0143 | 0.220 |
| Social withdrawal | -0.0103 | -0.0406 | 0.0201 | 0.505 |
| Addictive behaviours | 0.0219 | -0.0126 | 0.0564 | 0.213 |

**Supplementary Table 2 Statistical values of the associations between abstraction ability and psychopathology using the preregistered discrete metric.**

*Based on the results without the bootstrap method. Note that the p-value is not adjusted for the purpose of the reference.*

| | Coefficient | CI lower | CI upper | *p*-value |
|---|---|---|---|---|
| ADHD | -0.1314 | -0.2559 | -0.0069 | 0.039 |
| Psychological distress | -0.1221 | -0.2509 | 0.0067 | 0.063 |
| Obsessive-compulsive disorder | 0.0509 | -0.0782 | 0.1801 | 0.438 |
| Impulsivity | -0.1475 | -0.2739 | -0.0211 | 0.022 |
| Social anxiety | -0.0640 | -0.1897 | 0.0617 | 0.317 |
| Alcohol-related disorder | -0.1097 | -0.2316 | 0.0122 | 0.077 |
| Gaming disorder | -0.0424 | -0.1705 | 0.0856 | 0.515 |
| Nicotine-related disorder | -0.1414 | -0.2641 | -0.0187 | 0.024 |
| Compulsive hypersensitivity | 0.1238 | -0.0807 | 0.3283 | 0.234 |
| Social withdrawal | -0.1065 | -0.2683 | 0.0554 | 0.196 |
| Addictive behaviours | -0.1108 | -0.2938 | 0.0721 | 0.234 |

**Supplementary Table 3 Statistical values of the associations between the averaged confidence and psychopathology.**

*Based on the results without the bootstrap method. Note that the p-value is not adjusted for the purpose of the reference.*

| | Coefficient | CI lower | CI upper | *p*-value |
|---|---|---|---|---|
| ADHD | -0.0024 | -0.0115 | 0.0066 | 0.596 |
| Psychological distress | -0.0023 | -0.0117 | 0.0070 | 0.620 |
| Obsessive-compulsive disorder | -0.0168 | -0.0258 | -0.0077 | 0.000 |
| Impulsivity | 0.0045 | -0.0047 | 0.0137 | 0.334 |
| Social anxiety | 0.0045 | -0.0046 | 0.0135 | 0.334 |
| Alcohol-related disorder | -0.0086 | -0.0174 | 0.0002 | 0.054 |
| Gaming disorder | -0.0115 | -0.0207 | -0.0024 | 0.014 |
| Nicotine-related disorder | -0.0033 | -0.0122 | 0.0056 | 0.467 |
| Compulsive hypersensitivity | -0.0205 | -0.0347 | -0.0063 | 0.005 |
| Social withdrawal | 0.0214 | 0.0102 | 0.0326 | 0.000 |
| Addictive behaviours | -0.0083 | -0.0211 | 0.0045 | 0.203 |

**Supplementary Table 4 Statistical values of the associations between the metacognitive sensitivity and psychopathology.**

*Based on the results without the bootstrap method. Note that the p-value is not adjusted for the purpose of the reference.*

| | Coefficient | CI lower | CI upper | $p_{boot}$ |
|---|---|---|---|---|
| ADHD | -0.0173 | -0.0300 | -0.0039 | 0.012 |
| Psychological distress | -0.0201 | -0.0339 | -0.0057 | 0.006 |
| Obsessive-compulsive disorder | -0.0256 | -0.0394 | -0.0119 | 0.000 |
| Impulsivity | -0.0117 | -0.0271 | 0.0041 | 0.147 |
| Social anxiety | -0.0069 | -0.0223 | 0.0081 | 0.377 |
| Alcohol-related disorder | -0.0145 | -0.0299 | 0.0009 | 0.065 |
| Gaming disorder | -0.0099 | -0.0228 | 0.0040 | 0.153 |
| Nicotine-related disorder | -0.0136 | -0.0281 | 0.0016 | 0.079 |
| Compulsive hypersensitivity | -0.0372 | -0.0617 | -0.0118 | 0.003 |
| Social withdrawal | 0.0119 | -0.0077 | 0.0314 | 0.230 |
| Addictive behaviours | 0.0061 | -0.0150 | 0.0279 | 0.592 |

**Supplementary Table 5 Statistical values of the associations between abstraction ability and psychopathology with the bootstrap method.**

*Note that the p-value is not adjusted for the purpose of the reference.*

| | Coefficient | CI lower | CI upper | $p_{boot}$ |
|---|---|---|---|---|
| ADHD | -0.1296 | -0.2523 | -0.0023 | 0.048 |
| Psychological distress | -0.1206 | -0.2499 | 0.0078 | 0.068 |
| Obsessive-compulsive disorder | 0.0510 | -0.0811 | 0.1893 | 0.456 |
| Impulsivity | -0.1458 | -0.2653 | -0.0179 | 0.028 |
| Social anxiety | -0.0617 | -0.1902 | 0.0679 | 0.334 |
| Alcohol-related disorder | -0.1103 | -0.2287 | 0.0097 | 0.070 |
| Gaming disorder | -0.0398 | -0.1668 | 0.0958 | 0.548 |
| Nicotine-related disorder | -0.1439 | -0.2644 | -0.0254 | 0.019 |
| Compulsive hypersensitivity | 0.1199 | -0.0955 | 0.3292 | 0.263 |
| Social withdrawal | -0.1021 | -0.2708 | 0.0634 | 0.231 |
| Addictive behaviours | -0.1068 | -0.2973 | 0.0845 | 0.270 |

**Supplementary Table 6 Statistical values of the associations between the averaged confidence and psychopathology with the bootstrap method.**

*Note that the p-value is not adjusted for the purpose of the reference.*

| | Coefficient | CI lower | CI upper | $p_{boot}$ |
|---|---|---|---|---|
| ADHD | -0.0023 | -0.0115 | 0.0066 | 0.618 |
| Psychological distress | -0.0025 | -0.0113 | 0.0067 | 0.576 |
| Obsessive-compulsive disorder | -0.0168 | -0.0254 | -0.0079 | 0.000 |
| Impulsivity | 0.0045 | -0.0045 | 0.0139 | 0.335 |
| Social anxiety | 0.0043 | -0.0056 | 0.0148 | 0.422 |
| Alcohol-related disorder | -0.0087 | -0.0169 | -0.0005 | 0.038 |
| Gaming disorder | -0.0115 | -0.0196 | -0.0032 | 0.008 |
| Nicotine-related disorder | -0.0035 | -0.0125 | 0.0059 | 0.451 |
| Compulsive hypersensitivity | -0.0201 | -0.0361 | -0.0050 | 0.005 |
| Social withdrawal | 0.0211 | 0.0077 | 0.0350 | 0.002 |
| Addictive behaviours | -0.0084 | -0.0205 | 0.0040 | 0.185 |

**Supplementary Table 7 Statistical values of the associations between the metacognitive sensitivity and psychopathology with the bootstrap method.**

*Note that the p-value is not adjusted for the purpose of the reference.*

| | $\chi^2$ | *p*-value |
|---|---|---|
| ADHD | 0.009 | 0.923 |
| Psychological distress | 1.372 | 0.241 |
| Obsessive-compulsive disorder | 0.046 | 0.830 |
| Impulsivity | 2.000 | 0.157 |
| Social anxiety | 1.863 | 0.172 |
| Alcohol-related disorder | 0.001 | 0.971 |
| Gaming disorder | 0.715 | 0.398 |
| Nicotine-related disorder | 0.041 | 0.840 |
| Transdiagnostic model | 4.390 | 0.222 |

**Supplementary Table 8 Results of likelihood ratio tests to compare models**

*We compared simple models with fixed variables (i.e., psychopathology + metacognitive sensitivity) and models including interactions between metacognitive sensitivity and psychopathology. Based on the results without the bootstrap method.*
